# Evaluating the Clinical Utility of Artificial Intelligence Assistance and its Explanation on Glioma Grading Task

**DOI:** 10.1101/2022.12.07.22282726

**Authors:** Weina Jin, Mostafa Fatehi, Ru Guo, Ghassan Hamarneh

**Affiliations:** School of Computing Science, Simon Fraser University, Burnaby, Canada; Division of Neurosurgery, The University of British Columbia, Vancouver, Canada

**Author notes:** Corresponding author. 8888 University Dr, Burnaby, BC, Canada. V5A 1S6.

**Keywords:** Artificial Intelligence, Neuro-Imaging, Neurosurgery, Explainable Artificial Intelligence, Clinical Study

## Abstract

**Background:** As a fast-advancing technology, artificial intelligence (AI) has considerable potential to assist physicians in various clinical tasks from disease identification to lesion segmentation. Despite much research, AI has not yet been applied to neurooncological imaging in a clinically meaningful way. To bridge the clinical implementation gap of AI in neuro-oncological settings, we conducted a clinical user-based evaluation, analogous to the phase II clinical trial, to evaluate the utility of AI for diagnostic predictions and the value of AI explanations on the glioma grading task.

**Method:** Using the publicly-available BraTS dataset, we trained an AI model of 88.0% accuracy on the glioma grading task. We selected the SmoothGrad explainable AI Weina Jin and Mostafa Fatehi are co-first authors.

algorithm based on the computational evaluation regarding explanation truthfulness among a candidate of 16 commonly-used algorithms. SmoothGrad could explain the AI model’s prediction using a heatmap overlaid on the MRI to highlight important regions for AI prediction. The evaluation is an online survey wherein the AI prediction and explanation are embedded. Each of the 35 neurosurgeon participants read 25 brain MRI scans of patients with gliomas, and gave their judgment on the glioma grading without and with the assistance of AI’s prediction and explanation.

**Result:** Compared to the average accuracy of 82.5 *±* 8.7% when physicians perform the task alone, physicians’ task performance increased to 87.7 *±* 7.3% with statistical significance (*p*-value = 0.002) when assisted by AI prediction, and remained at almost the same level of 88.5 *±* 7.0% (*p*-value = 0.35) with the additional AI explanation assistance.

**Conclusion:** The evaluation shows the clinical utility of AI to assist physicians on the glioma grading task. It also reveals the limitations of applying existing AI explanation techniques in clinical settings.

**Key points:**

1. Phase II evaluation with 35 neurosurgeons on the clinical utility of AI and its explanation
2. AI prediction assistance improved physicians’ performance on the glioma grading task
3. Additional AI explanation assistance did not yield a performance boost

**Importance of the study:** This study is the first phase II AI clinical evaluation in neuro-oncology. Evaluating AI is a prerequisite for its clinical deployment. The four phases of AI clinical evaluation are analogous to the four phases of clinical trials. Prior works that apply AI in neurooncology utilize phase I algorithmic evaluation, which do not reflect how AI can be used in clinical settings to support physician decision making.

To bridge the research gap, we conducted the first clinical evaluation to assess the joint neurosurgeon-AI task performance. The evaluation also includes AI explanation as an indispensable feature for AI clinical deployment. Results from quantitative and qualitative data analysis are presented for a detailed examination of the clinical utility of AI and its explanation.

## 1 Introduction

Artificial intelligence (AI) and machine learning technologies have transformative potential in medicine, as evidenced by the ever-increasing research advances in medical AI in recent years [35, 33]. Using AI in medicine has the potential to support decision-making processes for healthcare providers and improve patient care. This is largely due to AI’s predictive capability to learn to recognize patterns from raw and high-dimensional data, such as medical images, electronic health records, and genomic data. In neuro-oncological settings, AI and machine learning have been studied in a wide range of applications, including identifying the grading and genetic mutations of brain tumors [25, 14, 13], predicting patients’ prognosis [47, 31], segmenting tumors based on magnetic resonance imaging (MRI) [36], triaging patients based on computed tomography (CT) scans [42], and discovering radiomics and radiogenomics for brain tumors [39]. Elsewhere, there are emerging cases of deploying AI in routine neuro-radiology practice, such as the AI-based RAPID software to detect vessel occlusion and triage stroke patients based on CT angiography [1].

Despite the above research advances, the widespread implementation of AI faces considerable challenges in translation from bench to bedside [21, 27]. A prerequisite to clinical deployment is bridging the AI evaluation gap between existing algorithmic evaluations and desired clinical evaluations. As with any new medical intervention, AI needs to undergo rigorous evaluation prior to its clinical implementation. Jin et al. [25] previously proposed four phases of clinical utility evaluation for AI in neuro-oncology, analogous to the conventional four-phases of clinical trials for drugs or medical devices: **phase I** is the algorithmic evaluation that evaluates the performance of AI model alone on unseen test data; From phase II and above, all evaluations include clinical users; **phase II** evaluates the primary efficacy of AI assistance on joint clinical user-AI task performance, conducted in *experimental* settings on *simulated* tasks; **phase III** further confirms the efficiency of the AI assistance on joint clinical user-AI task performance, conducted on a larger scale randomized controlled trial (RCT) in *clinical* settings on *real-world* tasks; and **phase IV** is for post-marketing software support and surveillance. Existing work in AI in neuro-oncology [13, 14, 47, 31, 36] has generally conducted phase I algorithmic evaluation, which can not reflect AI’s clinical utility in assisting clinical users in clinical workflow. Phase II clinical evaluation of AI has been conducted in other specialties such as orthopedics [9], psychiatry [23], and ophthalmology [37]. Phase III RCT clinical evaluation of AI in clinical settings had been conducted in specialties such as gastroenterology [45], and results showed a large variation of AI’s clinical utility [48, 40]. To the best of our knowledge, there are no phase II studies (and above) on the clinical utility of AI assistance in neuro-oncology, and this is the research gap we aim to bridge in this study.

In addition to the above AI evaluation gap, another significant hurdle for AI clinical implementation is the interpretability or explainability problem of AI. The state-of-theart AI models, namely deep neural networks, are black-box models, and their decision processes are incomprehensible even to AI engineers. This impedes the clinical use of AI, as clinical users will often require an explanation or justification from AI other than a mere prediction, due to the high-stakes nature of clinical decision-making [44]. Furthermore, explanations may enable physicians to identify potential errors of AI, and improve the joint physician-AI team performance [43, 46, 12, 6]. Therefore, we leverage the latest technical advance in explainable AI (XAI) as a feature of the AI system in our clinical evaluation study.

In this work, we recruited physicians and conducted a phase II clinical evaluation of AI in an experimental setting on a simulated clinical task based on brain MRI: classifying a glioma case into a glioblastoma (GBM, WHO grade IV), or a WHO grade II or III glioma. This is a clinically relevant question that helps guide subsequent management decisions. Tumor grading is also a routine and ubiquitous task in neuro-oncological settings and is commensurate with our participants’ knowledge in neuro-oncology. Ultimately, other tumor genetic characteristics, such as isocitrate dehydrogenase (IDH) mutation status and O6-methylguanine-DNA methyltransferase (MGMT) methylation status, are critical for treatment and prognostication, but are more challenging for clinicians to predict from imaging studies. Thus, in the present proof-of-concept study, we have focused on the task of differentiating GBM from grade II/III diffuse gliomas. Traditionally, clinicians have relied upon patient characteristics, image findings, along with neurological signs and symptoms to decide whether to proceed with an aggressive resection, perform a biopsy, or to continue with watchful waiting. The interpretation of imaging findings is contingent upon the neuro-radiologist’s and neurosurgeon’s experience, and this likely contributes to some of the heterogeneity seen in practice between clinicians who treat patients with gliomas [18]. A potential AI-based tool that can accurately predict tumor genetics and histologic grading would potentially not only decrease the heterogeneity in management, but also help guide biopsy plans and improve the ability to prognosticate outcomes.

In our evaluation, we conducted a nationwide clinical study in Canada on the glioma grading task. We recruited 35 neurosurgeons, each of whom read a set of 25 brain MRIs without and with AI assistance (glioma grade prediction and explanation). Results showed that physicians’ average task accuracy improved from 82.5% without AI assistance to 87.7% with the assistance of AI prediction (*p* = 0.002), and such accuracy did not change with the additional assistance of AI explanation (*p*-value = 0.35). The results confirm the effect of AI to enhance physicians’ clinical task performance in a simulated clinical setting. This is the first study in neuro-oncology to evaluate the clinical utility of AI assistance.

## 2 Study Material

### 2.1 MRI data

We used the publicly-available Multimodal Brain Tumor Segmentation (BraTS) 2020 dataset [32, 5] in the glioma grading clinical study, as well as to train the AI model. The BraTS dataset contains routine clinically-acquired, pre-operative brain MRI scans from patients with glioma. The brain MRIs in BraTS dataset were obtained with different clinical protocols and various scanners from 19 institutions, including the publicly-available TCGA/TCIA repositories [4, 3]. Each MRI scan consists of four MRI pulse sequences of T1-weighted, T1-weighted contrast enhancing (T1C), T2-weighted, and T2 Fluid Attenuated Inversion Recovery (FLAIR). MRIs in the BraTS dataset were pre-processed images with the pre-processing steps of co-registration to the same T1 anatomic template, resampling to 1*mm*^3^ voxel resolution, and skull-stripping. The pre-processing methods are detailed in Bakas et al. [5]. Each MRI scan is associated with a tumor grade label of a GBM or grade II/III glioma, which was pathologically confirmed. The total number of MRI cases are 369, of which 76 are grade II/III glioma cases, and 293 are GBM cases.

### 2.2 AI model and algorithmic evaluation on glioma grading task

We trained an AI model using the BraTS 2020 dataset to grade glioma MRIs. The AI model receives an MRI input and outputs a glioma grade of either a GBM or a grade II/III glioma. The model architecture is a VGG-like [38] three-dimensional (3D) convolutional neural network (CNN), with six 3D CNN layers connected to two fully connected layers. We stratified split the BraTS dataset into 65% training (239 cases), 15% validation (56 cases), and 20% (74 cases) hold-out test set by keeping the same grade II/III : GBM ratio in each set. There were no patient’s ID overlapping across the three datasets. The training, validation, and test accuracies of the AI model are 80.28%, 92.86%, and 90.54%, respectively. The fine-grained model performance metrics are in Supplemental S2 Fig. 1, which was also shown to participants in the clinical study.

From the test set, we sampled a subset of 25 MRIs as the clinical test subset used in the glioma grading clinical study. We sampled the subset by keeping the same ratio of the correctly/incorrectly predicted grade II/III glioma or GBM as the confusion matrix of model performance in Supplemental S2 Fig. 1. This is to keep an equivalent performance of the AI model on the test set and the clinical test subset. In the clinical test subset, there are 7 cases of grade II/III glioma, and 18 cases of GBM. The AI model has an accuracy of 88.00% on the clinical test subset.

### 2.3 Generating and selecting the optimal AI explanation

The AI model we trained to grade glioma is a black-box CNN model. To explain the model decisions to physicians, we applied post-hoc XAI algorithms that act as a surrogate model to approximate the black-box AI model by probing the model parameters and/or input-output pairs. From a candidate list of 16 post-hoc XAI algorithms that can generate feature attribution map or heatmap (named as color map in the user study) to explain the important image regions for model prediction, we selected SmoothGrad [41], which is most truthful to the AI model decision process [26]. The evaluation method and result are detailed in Supplemental S2.

## 3 Method

### 3.1 Participants

We recruited physicians to evaluate their clinical task performance without and with AI assistance. The inclusion criteria for the study participants were: the participant must hold an MD or equivalent; and must be a consultant neurosurgeon, radiologist, or neuroradiologist, or a trainee in neurosurgery, radiology, or neuro-radiology. Since the study was conducted anonymously as an online survey, two stages of eligibility screening were conducted: one was conducted at the beginning of the online survey, where participants were filtered by their answers to the questions about their roles in medical practice and their medical specialty. The other was conducted using a post-survey screening process to filter out responses that do not meet the inclusion criteria due to random guess or lack of required expertise in neuro-oncological MRI interpretation. We did so by only including participants whose task accuracy when performing the grading task alone was above 0.55. The accuracy threshold was set to be slightly higher than the random guess accuracy of 0.5. We used convenience sampling and recruited participants by directly contacting the researchers’ national-wide clinical research network. The recruitment period was from October 2021 to February 2022.

### 3.2 Study design and procedure

We designed a pre-post clinical study to examine the clinical utility of AI system regarding its benefit to physicians’ task performance. The study consisted of an online glioma grading survey (30-40 minutes) and an optional remote interview (20-30 minutes). Participants provided separate consents for the survey and the interview.

The online survey is the main part of the study, where participants read a set of 25 MRIs without and with the assistance of AI prediction and its explanation. The MRIs were sampled from the BraTS 2020 dataset as described in Section 2.1. The sequence in which MRIs were shown was randomized for each participant to avoid bias due to MRI reading order. Participants were first introduced to the AI system and its performance on the test set. Then they began the MRI reading task. For each MRI, the participants first gave their own judgment. Then the AI’s prediction was revealed to them, and they were asked to give their current and possibly updated judgment of the glioma grade. The AI prediction was shown only after participants gave their own judgment. Next, participants were asked about their willingness to check AI’s explanation of how it arrived at the prediction, and were shown a heatmap explanation from AI with important regions for AI’s prediction highlighted (an example image is shown in Supplemental S2 Fig.2). The MRI and heatmap explanation were both 3D images shown in video format, in which participants could control the video play to view different MRI slices and corresponding heatmap explanation. After that, participants were asked to provide their final (again, possibly modified) judgment on the glioma grade, and evaluate the agreement between their own clinical judgment and heatmap on an 11-point scale from 0 to 10.

We also asked participants to rate, on an 11-point scale, their trust level in the AI system and willingness to incorporate this AI’s suggestions into routine clinical practice. The two questions were asked at three time points: at the beginning of the survey before exposure to any information on the AI system as baseline, after viewing AI’s performance metrics (Supplemental S2 Fig. 1) on the test set, and after using AI with its prediction and explanation assistance for the 25 MRIs. The survey ends with the question on the ranking of possible goal(s) of checking AI explanations, and a short demographic questionnaire on the participant’s medical experience, familiarity with AI, attitude towards AI, age, and gender. The full survey content is in Supplemental S3.

After completing the online survey, participants were given monetary compensation ($50 CAD gift card) as appreciation for their time and effort. The participant could choose to participate in an optional remote interview. In it, participants talked about their user experience and commented on the AI system. Participants provided additional consent prior to the interview. The remote interview sessions were videoor audiorecorded for qualitative data analysis. The study was approved by the Research Ethics Board of Simon Fraser University (Ethics number: H20-03588).

### 3.3 Statistical Analysis

We conduct statistical analysis to test for the following null hypotheses:

1. There are no differences in physicians’ accuracies on the glioma grading task, across the three conditions: 1) Physician performing the task alone, denoted as <u>DR</u>; 2) Physician performing the task with the assistance of AI prediction, denoted as <u>DR+AI</u>; 3) Physician performing the task with the assistance of AI prediction and explanation, denoted as <u>DR+XAI</u>.
2. There are no differences in the physicians’ trust level across the three time points: 1) Initial baseline without knowing any information from AI; 2) After viewing AI performance metrics; and 3) After using AI with its prediction and explanation assistance for the 25 MRIs.
3. There are no differences in the physicians’ willingness to use AI across three time points: 1) Initial baseline without knowing any information from AI; 2) After viewing AI performance metrics; and 3) After using AI with its prediction and explanation assistance for the 25 MRIs.

To test the above three hypotheses, a one-way analysis of variance (ANOVA) with repeated measures is performed when data fulfill the assumptions of normality and sphericity. We use Shapiro-Wilk test of normality and Mauchly’s test for sphericity to test the assumptions for ANOVA. If the null hypothesis is rejected, a post-hoc analysis is conducted using Tukey’s HSD (honestly significant difference) test when data met the assumption of homogeneity of variances. Otherwise, if assumptions for ANOVA are violated, we use the non-parametric Friedman test, and a post-hoc analysis using Wilcoxon signed-rank test with Bonferroni correction.

Additionally, the Spearman correlation coefficient is used to measure the association between two continuous variables; and the chi-square test of independence is conducted to test the association between two categorical variables. Unless otherwise stated, we use a significance level *α* = 0.05. The statistical analysis was performed using Python statistical package SciPy^1^ and Pingouin^2^. We make the survey data and analysis code available in Supplemental S4 for reproducibility.

#### 3.3.1 A pilot study to estimate sample size

Before launching the formal national study, we conducted a pilot study to iterate the survey content and estimate the sample size. Six neurosurgical residents were recruited in the pilot study. With a two-sided test size of 5% and a power of 90%, based on the effect size of 1.3 between <u>DR</u> and <u>DR+AI</u>, the estimated sample size is 13.

### 3.4 Presentation of results

In the manuscript, we report the quantitative analysis of the survey data, and provide the full results of the qualitative data analysis in Supplemental S1. The qualitative data are from the interview and free-text input in the survey. We discuss findings from both quantitative and qualitative data in the Discussion Section 5. We number the participants with N1, N2, … when directly quoting their words.

## 4 Result

### 4.1 Participants

A total of 35 participants met the inclusion criteria and were enrolled in the study. The recruitment rate was 14.8% (35 out of 236 eligible participants contacted). Among them, 29 participants completed the survey, while 6 participants dropped out without completing the survey (their numbers of completed MRI interpretation cases were: 2, 3, 3, 3, 8, and 17, respectively). In addition, five participants took the interview to provide qualitative comments on the use of the AI system in the survey. The recorded selfreported demographic data of the participants were: female: male = 7:19; age: 34.7 *±* 8.2 (mean ± std); all participants were from the neurosurgery specialty, and their positions were: 12 attending neurosurgeons, 2 neurosurgical fellows, and 21 neurosurgical residents. Their years of practicing medicine were 9.8 *±* 9.1; and their years of practicing neurosurgery were 7.1 *±* 6.5. Figure 1 summarizes their familiarity with AI and their attitude towards AI. Most participants (96%) are familiar with AI technologies, e.g. they had heard of AI, or had used it at work or in their daily lives. Over 2/3 of participants had a positive attitude towards AI, whereas the rest had a skeptical or neutral attitude. The detailed demographics are listed in Supplemental S2 Table 1.

**Figure 1:**
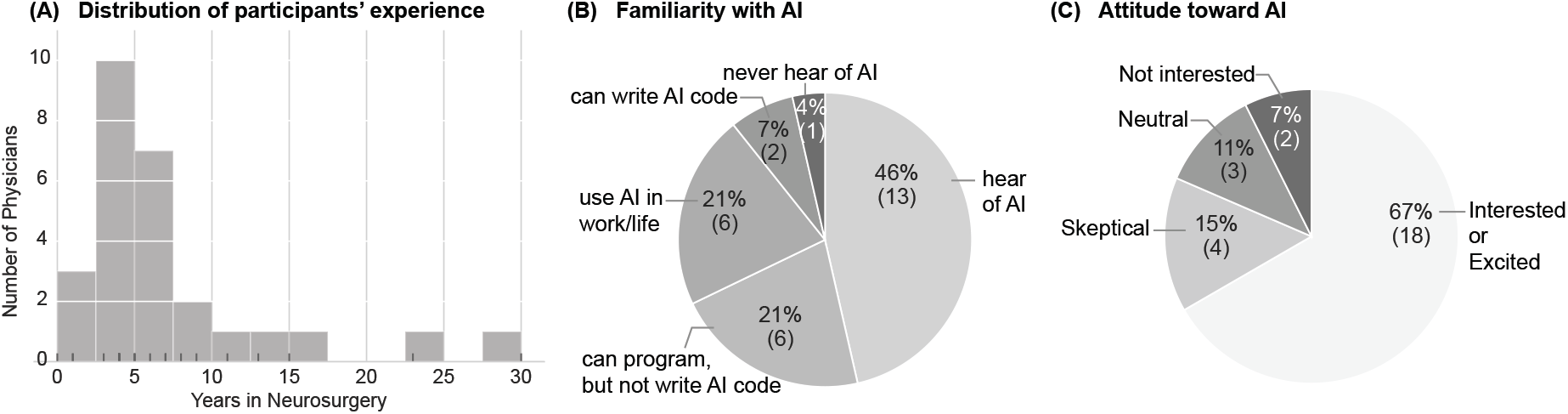
Participants’ demographic information. Panel (A) shows the distribution of participants’ years of neurosurgical practice; The sticks on the *x*-axis show each participant’s years of practice. Panel (B) shows participants’ familiarity with AI, and panel (C) shows their attitude towards AI, with the percentage and number of participants (in parentheses) for each catalog indicated.

### 4.2 Physicians’ task performance in three decision-support conditions

A total of 2279 glioma grading decisions were collected for the three decision-support conditions of 1) <u>DR</u>: physician performing the task alone (761 decisions); 2) <u>DR+AI</u>: physician performing the task with AI prediction assistance (759 decisions); and 3) <u>DR+XAI</u>: physician performing the task with AI prediction and explanation assistance (759 decisions). Participants’ average task accuracies for the three conditions were: <u>DR</u>: 82.49 *±* 8.69% (mean*±*std), <u>DR+AI</u>: 87.70 *±* 7.33%, and <u>DR+XAI</u>: 88.52 *±* 7.02%. The descriptive statistics of participants’ task accuracy for the three conditions are in Table 2.

**Table 1:** List of variables collected in the survey, and their corresponding survey questions. For the trust scale, we post-processed the responses by adding 5 to all responses, so that the scale range is from 0 to 10. In the following text, we underline the variable names listed in the table. Variables above the double horizontal line were asked for each MRI case, and the ones below were only asked once or several times at different time points.

| Variable name | Survey question | Survey question options |
| --- | --- | --- |
| <u>DR</u> | What grade of glioma would you predict this MRI to be? | • Grade II/III glioma<br>• Grade IV glioma |
| <u>DR+AI</u> | After viewing AI’s suggestion, what is your current judgment on the tumor grade? | ditto |
| <u>DR+XAI</u> | After viewing AI’s explanation, what is your final judgment on the tumor grade? | ditto |
| <u>Need explanation</u> | Would you like to check the explanation from AI for this MRI? | Yes/No option |
| <u>Explanation quality</u> | How closely does the highlighted area of the color map match with your clinical judgment? | [0-10 scale]<br>• 0, Not close at all<br>• 5, Somewhat close<br>• 10, Very close |
| <u>Trust</u> | What is your trust level in this AI model? | [Scale from -5 to 5]<br>• -5, Totally distrust the AI<br>• 0, Neutral, neither distrust nor trust<br>• 5, Totally trust the AI |
| <u>Willingness to use AI</u> | How likely will you incorporate this AI’s suggestions into your routine clinical practices, such as diagnosis, prognosis, and medical management? | [0-10 scale]<br>• 0, Not likely<br>• 5, Somewhat likely<br>• 10, Very likely |
| <u>Explanation goal</u> | When are you most likely to check those color map explanations from AI? | Select and rank from a set of 15 predefined and a self-filled options |

**Table 2:**
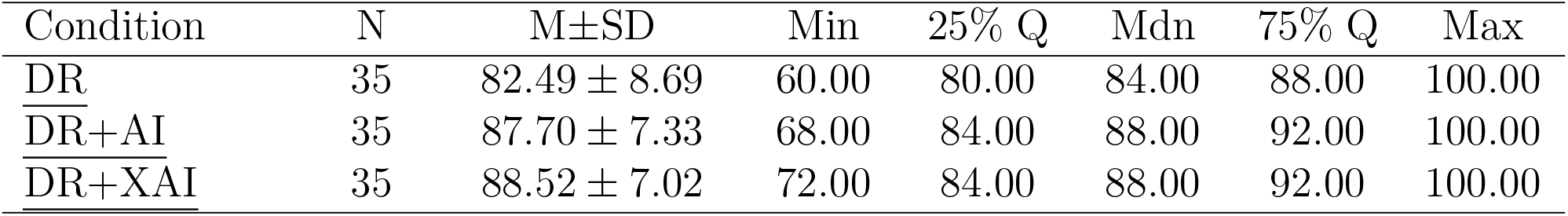
Descriptive statistics for all participants’ task performance accuracy (%). N number of participants, M - mean, SD - standard deviation, Q - quantile, Mdn - median.

All data passed the sphericity assumption test for ANOVA, but the data on <u>DR</u> condition did not pass the normality assumption. Therefore, we used the non-parametric Friedman test instead, and results showed a statistically significant difference in task accuracies among the three conditions, 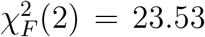, *p <* 0.001. We then conducted post-hoc analysis using Wilcoxon signed-rank tests with Bonferroni correction. Results showed that the <u>DR+AI</u> condition had a statistically higher accuracy compared to the <u>DR</u> condition (*Z* = 9.0, *p* = 0.002); similarly, the <u>DR+XAI</u> condition had a statistically higher accuracy compared to the <u>DR</u> condition (*Z* = 3.0, *p* = 0.0004). However, the accuracy values across the <u>DR+AI</u> and <u>DR+XAI</u> conditions did not show statistically significant difference (*Z* = 15.5, *p* = 0.35) (Fig. 2). We also calculated the effect size using common language effect size, and results showed a physician has a probability of 67.2% of having a higher accuracy when assisted by AI prediction (<u>DR+AI</u>) than performing the task alone (<u>DR</u>), a probability of 71.0% of having a higher accuracy when assisted by AI prediction and explanation (<u>DR+XAI</u>) than performing the task alone (<u>DR</u>), but only a probability of 53.6% of having a higher accuracy when assisted by AI prediction and explanation (<u>DR+XAI</u>) than assisted by AI prediction alone (<u>DR+AI</u>). In addition to the above result analysis using the performance accuracy metric, we also report the results and statistical tests regarding other performance metrics, including accuracy, sensitivity, specificity, and F1 score in Supplemental S2 Table 3.

**Figure 2:**
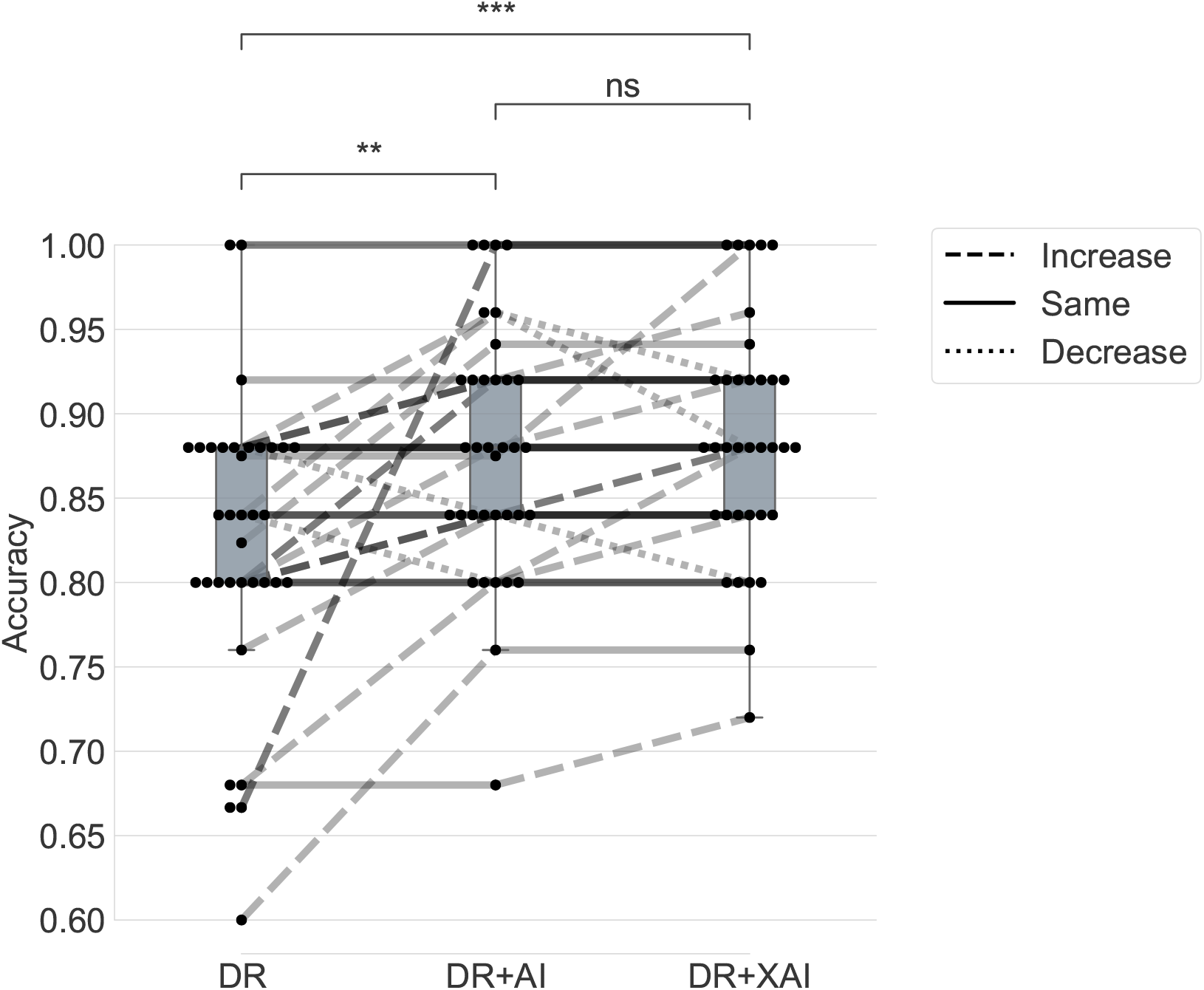
Participants’ task performance on glioma grading in three conditions: 1) <u>DR</u>: Physicians performing the task alone; 2) <u>DR+AI</u>: Physicians performing the task with AI assistance (with predictions from AI); 3) <u>DR+XAI</u>: Physician performing the task with XAI assistance (with predictions and explanations from AI). We show box plots for the three conditions, with lines and dots indicating the change of performance for each participant. The dots indicate individual participants’ performances. The lines connecting dots indicate a participant’s performance change in between different conditions, with a dashed line indicating an increment, a solid line indicating no change, and a dotted line indicating a decrement. ns: *p >* 0.05, *\*\** : 0.001 *≤ p ≤* 0.01, *\* * \** : 0.0001 *≤ p ≤* 0.001.

### 4.3 Decision agreement and decision change patterns

We analyzed the decision agreement and decision change for each fine-grained decision in the three conditions (<u>DR</u>, <u>DR+AI</u>, and <u>DR+XAI</u>), and visualized such patterns as decision change stream plot in Fig. 3. The subgroup analysis for attending and resident + fellow physician subgroups showed similar patterns (Supplemental S2 Fig.7).

**Figure 3:**
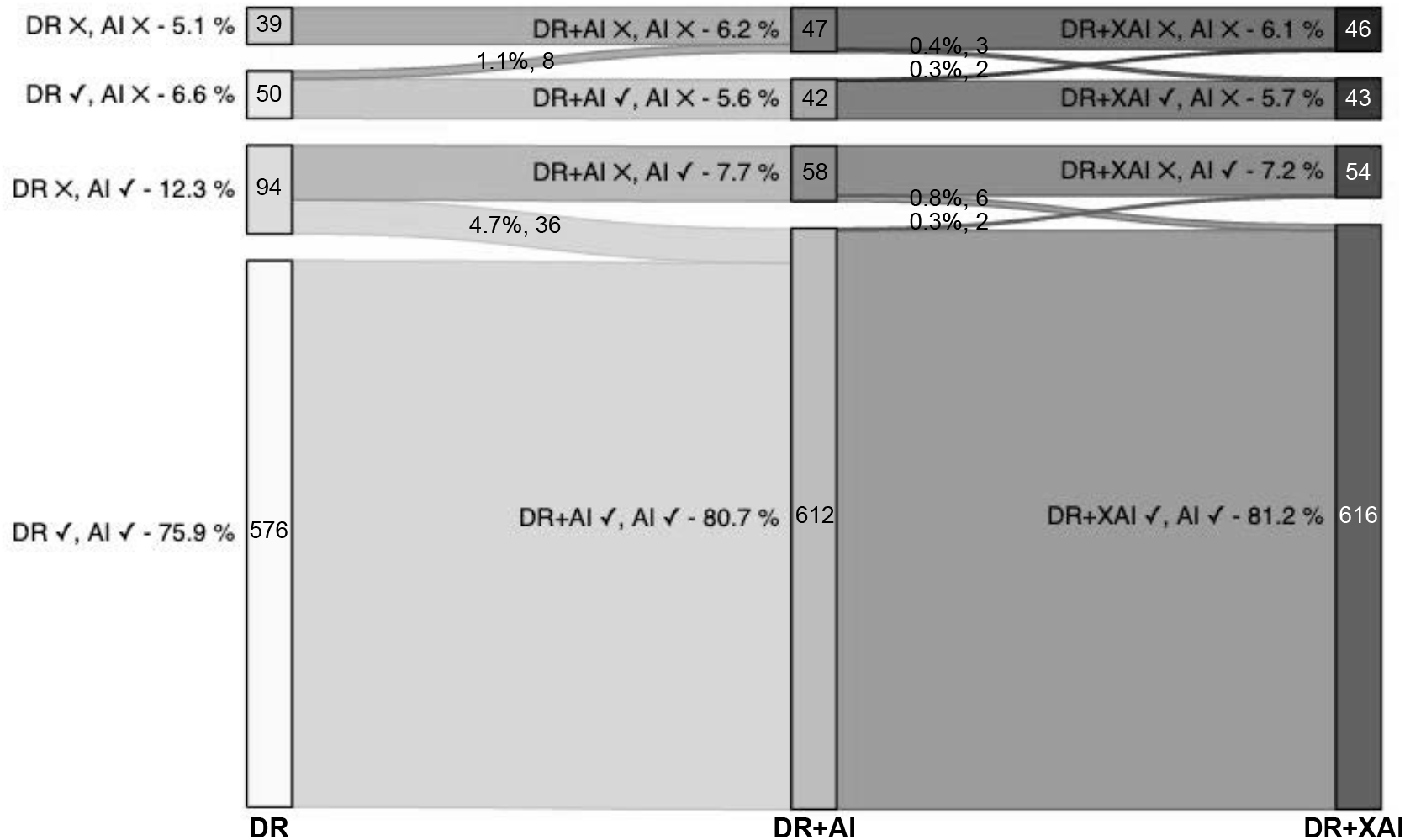
Participants’ decision change stream plot for each error category of all participants. The three columns represents the three conditions of <u>DR</u>, <u>DR+AI</u>, and <u>DR+XAI</u>, respectively. The four rectangles within each column records the number and percentages of cases when the doctor and the AI’s decision where correct (✓) or not (*×*), e.g., the decision agreement tally is reported in the first and fourth rectangles, reflecting when the doctor and AI made the same decisions (whether both are correct or incorrect). The total number of decisions is 759 for each column.

For the decision agreement pattern, as shown in Fig. 3, as a baseline when physicians performed the task alone (<u>DR</u>), physicians and AI’s decisions agree with each other in 81.0% (615/759) decisions. The decision agreement increased to 86.8% (659/759) when physicians were assisted by AI’s prediction (<u>DR+AI</u>), and further increased to 87.2% (662/759) when physicians were assisted by both AI’s prediction and explanation (<u>DR+XAI</u>).

For physicians’ decision change pattern during AI assistance, as shown in Fig. 3, with assistance by AI’s prediction (<u>DR+AI</u> condition), physicians changed 5.8% (44/759) of their original decision to AI’s prediction, and such decision change occurred only during decision disagreement between AI’s prediction and physicians’ decision. Among these decision change cases, 81.8% (36/44) of the change where correct changes, i.e., resulted in a corrected decision (i.e., updated judgment matched the ground truth diagnosis), and the remaining 18.2% (8/44) were incorrect changes, i.e., leading to an erroneous decision. With further assistance by AI’s explanation (<u>DR+XAI</u> condition), physicians changed 1.7% (13/759) of their decisions, and such decision change occurred during both decision agreement (0.7%, 5/759) and disagreement (1.1%, 8/759) between AI’s prediction and physicians’ decision. Among them, 69.2% (9/13) changed correctly, and 30.8% (4/13) changed incorrectly.

### 4.4 Trust and willingness to use AI

We tested whether participants would calibrate their level of <u>trust</u> in the tested AI system and their <u>willingness to use AI</u> with the exposure to AI’s performance metrics and the actual AI usage experience. Participants’ level of <u>trust</u> in AI and <u>willingness to use AI</u> were recorded at three time points: 1) the initial baseline without knowing any information from AI; 2) after viewing AI’s performance metrics, and 3) after using AI’s predictions and explanations for the 25 MRIs. The descriptive statistics of the two variables at three time points are listed in Table 3. In addition, the two variables <u>trust</u> in AI and <u>willingness to use AI</u> are highly correlated, with a Spearman correlation coefficient of 0.70 (*p <* 0.001).

**Table 3:**
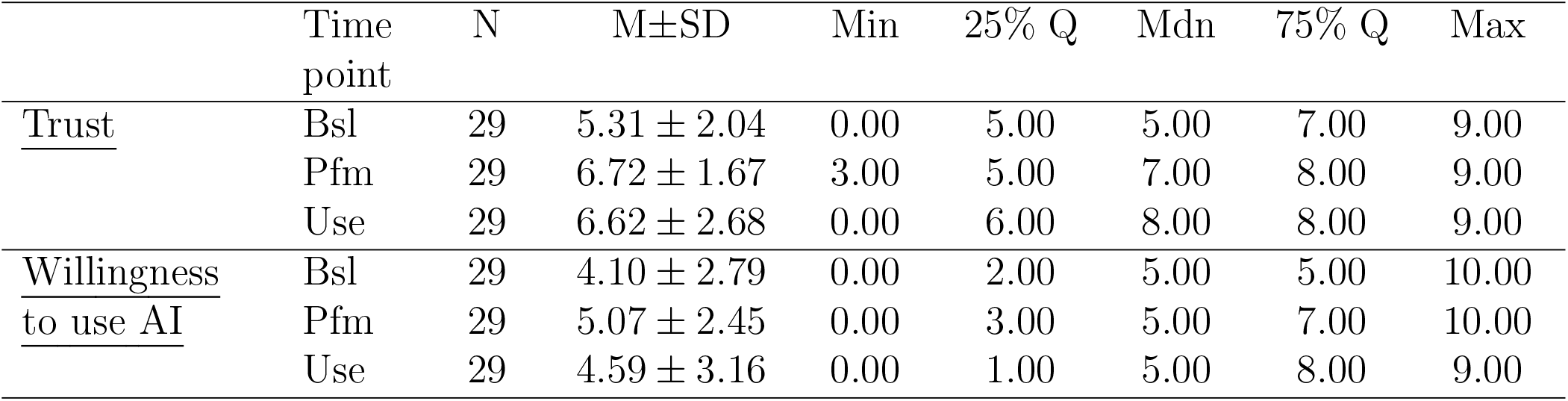
Descriptive statistics for participants’ trust and willingness to use AI. N - number of participants, M - mean, SD - standard deviation, Q - quantile, Mdn - median. The three time points are: 1) Bsl: the initial baseline without knowing any information from AI; 2) Pfm: after viewing AI performance metrics, and 3) Use: after using AI’s predictions and explanations for the 25 MRIs.

| | Time point | N | M±SD | Min | 25% Q | Mdn | 75% Q | Max |
| --- | --- | --- | --- | --- | --- | --- | --- | --- |
| <u>Trust</u> | Bsl | 29 | 5.31 ± 2.04 | 0.00 | 5.00 | 5.00 | 7.00 | 9.00 |
| | Pfm | 29 | 6.72 ± 1.67 | 3.00 | 5.00 | 7.00 | 8.00 | 9.00 |
| | Use | 29 | 6.62 ± 2.68 | 0.00 | 6.00 | 8.00 | 8.00 | 9.00 |
| <u>Willingness to use AI</u> | Bsl | 29 | 4.10 ± 2.79 | 0.00 | 2.00 | 5.00 | 5.00 | 10.00 |
| | Pfm | 29 | 5.07 ± 2.45 | 0.00 | 3.00 | 5.00 | 7.00 | 10.00 |
| | Use | 29 | 4.59 ± 3.16 | 0.00 | 1.00 | 5.00 | 8.00 | 9.00 |

Part of the <u>trust</u> and <u>willingness to use AI</u> data did not pass the sphericity and normality assumption test for ANOVA. Therefore, we used the non-parametric Friedman test instead. Results showed a statistically significant difference among the three time points for both <u>trust</u> in AI (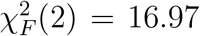, *p* = .0002), and <u>willingness to use AI</u> (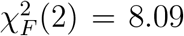, *p* = .018). We conducted post-hoc analysis using Wilcoxon signed-rank tests with Bonferroni correction to identify the statistically different pairs. For the level of <u>trust</u> in the AI system, participants rated a statistically higher trust after viewing AI’s performance metrics compared with the initial baseline (*Z* = 4.0, *p* = .0004); and a statistically higher trust after using AI’s predictions and explanations for the 25 MRIs compared with the initial baseline (*Z* = 58.0, *p* = .025); but there was no statistically difference between the trust level after viewing AI’s performance metrics, and after using AI’s predictions and explanations for the 25 MRIs. For the level of <u>willingness to use AI</u>, participants only rated a statistically higher willingness to use AI after viewing AI’s performance metrics compared with the initial baseline (*Z* = 29.5, *p* = .012); and the rest pairwise test did not show a statistically significant difference. The statistical test results are visualized in Fig. 4.

**Figure 4:**
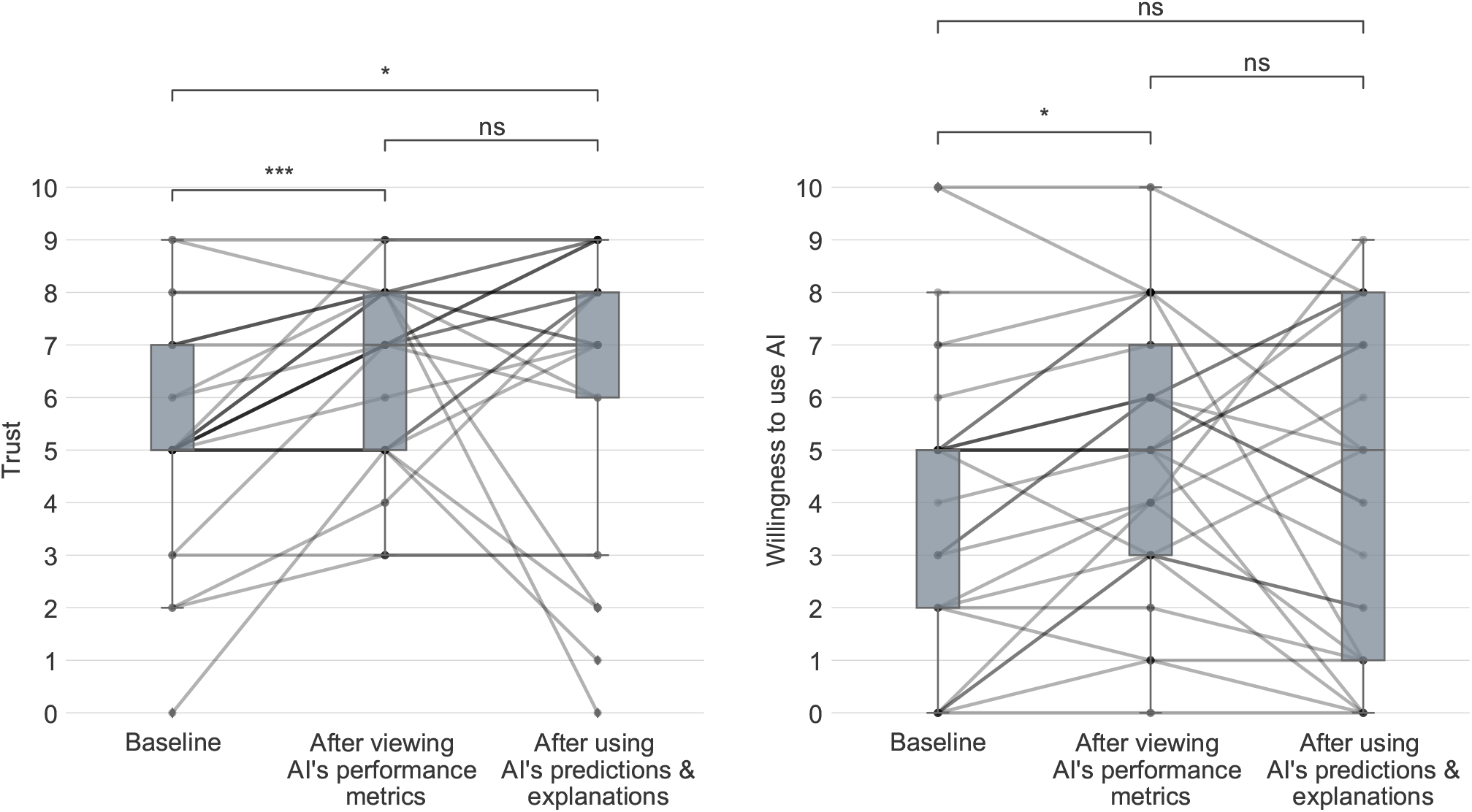
Box plots and changes of participants’ trust in the AI (left), and willingness to use AI (right) at the initial baseline, after viewing AI’s performance metrics, and after using the AI’s predictions and explanations. Both dependent variables are reported on a 0-10 point scale. The lines in between two time points indicate the change for each participant. ns: *p >* 0.05, *\** : 0.01 *≤ p ≤* 0.05, *\*\** : 0.001 *≤ p ≤* 0.01, *\* * \** : 0.0001 *≤ p ≤* 0.001.

### 4.5 Clinical usage scenarios for AI explanation

Physicians’ behavior of seeking explanation is usually to support their subsequent clinical sub-tasks. We summarized such potential <u>explanation goals</u> from literature [24], and asked participants to select and rank them. The results are shown in Table 4. The top-rated explanation goals are related to the critical nature of the clinical task, and the explanation is useful mainly to safeguard the clinical decision.

**Table 4:** Ranking of explanation goals. We report under heading “Selected times” the number of times each goal was selected by participants, and “Rankings” shows participants’ individual ranking (in bold) and times of such ranking (in parentheses).

| Explanation goal | Selected times | Rankings |
| --- | --- | --- |
| To build and calibrate my trust in this AI | 18 | **1** (7), **2** (4), **3** (3), **4** (3), **5** (1), |
| When I doubt about the prediction from AI | 15 | **1** (8), **2** (1), **3** (3), **4** (3), |
| To verify AI’s decisions | 16 | **1** (2), **2** (6), **3** (3), **4** (2), **5** (1), **6** (2), |
| To ensure the safety use of the AI | 11 | **1** (2), **2** (2), **3** (4), **4** (3), |
| For a difficult case, when I am not certain | 10 | **1** (1), **2** (3), **3** (1), **4** (2), **5** (2), **7** (1), |
| To learn from AI | 7 | **2** (1), **3** (1), **4** (2), **6** (2), **8** (1), |
| To improve my patients’ outcomes | 5 | **1** (3), **4** (1), **5** (1), |
| To ensure fairness and no biases in the AI model | 5 | **2** (1), **3** (2), **5** (1), **9** (1), |
| When I am trading off among multiple objectives for my patient | 2 | **2** (1), **3** (1), |
| To meet the ethical requirements | 2 | **2** (1), **5** (1), |
| To make Differential Diagnosis | 3 | **3** (1), **5** (1), **10** (1), |
| To make new medical discovery | 3 | **5** (1), **6** (1), **7** (1), |
| Before discussion with my colleagues | 1 | **5** (1), |
| To meet the legal requirements | 1 | **6** (1), |
| To generate report or patient chart | 0 |  |

In addition to the above general <u>explanation goal</u> in a clinical setting, for each MRI case, we asked a yes/no question on whether the participant needs to check the AI’s explanation (<u>need explanation</u>). We calculated each participant’s <u>need explanation</u> degree (0: no need for explanation, and 1: need for explanation for all cases) by the ratio of “yes” answers out of all the recorded responses among the 25 MRI cases. The trend of <u>need explanation</u> shows polarization, with 66% (23/35) of the participants had a <u>need explanation</u> degree of less than 0.3, and 20% (7/35) had a <u>need explanation</u> degree of above 0.7. In particular, 14% (5/35) participants completely did not need any explanation for all cases (<u>need explanation</u> degree = 0), and 9% (3/35) of participants needed explanation for every case (<u>need explanation</u> degree = 1). We further conducted a chi-square test of independence on whether <u>need explanation</u> is associated with decision agreement, and there is a significant relationship between the two variables, *χ*^2^(1) = 62.7, *p <* 0.001. When there is a decision disagreement between AI and physicians’ initial judgment, physicians are more likely to check the explanation.

For the quality of the specific explanation content, we collected 744 ratings on the heatmap <u>explanation quality</u> on a 0-10 point scale, and obtained an average quality rating of 6.12*±*2.92 (mean*±*std). Each heatmap explanation received ratings with large standard deviations ranging from 2.39 to 3.04. The rating for each of the 25 AI explanations is listed in Supplemental S2 Table 2, and we visualized the explanation with the highest and lowest average rating in Supplemental S2 Fig.2.

## 5 Discussion

### 5.1 The clinical utility of AI prediction

In our study focusing on the glioma grading clinical task, physician’s initial task performance was lower than AI’s performance. With AI assistance, physicians’ task performance significantly increased to be equivalent to AI’s performance. The result aligns with prior phase II clinical evaluation of AI on medical image analysis tasks to diagnose knee lesions [9], diabetic retinopathy [37], and pulmonary adenocarcinoma [29], where physicians exhibited better task performance with the assistance of a superior AI (the AI that outperformed physicians); but diverge from the similar phase II study in psychiatry on medical record data [23], where a superior AI assistance did not improve clinicians’ treatment selection accuracy. These divergent results indicate the variability in the effect of AI assistance in clinical settings, and suggest the importance of conducting clinical studies to validate the clinical utility of AI assistance on specific clinical tasks.

Our clinical study validates AI’s clinical utility as a physician performance booster in the glioma grading task. Such quantitative result are echoed by the qualitative results (Supplemental S1 Section 1) wherein physicians regard AI as a “second opinion” (N2), or “another level of evidence” (N5). Quantitative results showed that such performance improvement is more prominent for junior physicians (residents and fellows, Supplemental S2 Section 2.2). Qualitative results further showed junior physicians benefit from AI in time-sensitive cases and hard cases; and can improve junior physicians’ learning and problem-solving skills by “reaffirming what you’re learning” (N2).

By further analyzing the decision change pattern, the observed physicians’ task performance improvements with AI prediction assistant were mainly due to the fact that physicians’ decision patterns converges to be more similar to AI’s decision, as physicians’ only switched their decisions during decision disagreement (Fig. 3). The decision disagreement between physicians and AI caused physicians “to pause and then go through the images … to understand the disagreement” (N5). But since in this condition, physicians only assisted by AI prediction alone, they could not get access to more information to resolve disagreement, and one of the expected utilities of an AI explanation is to provide such information to facilitate physicians’ decision-making.

In addition, our study also inspected factors that influence physicians’ adoption of AI for decision support, including their trust and willingness to use AI. We noticed that after viewing AI’s performance metrics, physicians’ trust and willingness to use AI significantly increased compared to the baseline. However, when used the AI on 25 cases with the assistant of AI’s prediction and explanation, physicians’ trust and willingness to use AI diverged, and the average ratings of trust and willingness to use AI did not show statistical difference compared with the ones after only viewing AI’s performance metrics. This indicates physicians may perceive different messages from AI prediction and explanation information, and construct different mental models of AI [7] accordingly to calibrate their trust and willingness to use AI. Such hypothesis is evidenced by physicians’ positive and negative comments for AI’s explanation from qualitative data (detailed in Section 5.2). Furthermore, qualitative results showed that to establish trust, some physicians request more information beyond the model performance metrics alone, such as information on model confidence and dataset (Supplemental S1 Section 3.3).

### 5.2 The clinical utility of AI explanation

In the study, with the additional assistance of AI explanation, physicians’ task performance did not show statistical difference compared to the performance with AI prediction assistance only. The finding aligns with a similar phase II clinical study involving AI heatmap explanation in diabetic retinopathy [37], and other similar AI-supported decision-making experiments involving laypersons on an age prediction task [15], and on a criminal justice decision support [2], where presenting AI prediction alone would improve human accuracy, but there was no additional performance boost with the assistance of feature attribution (i.e. heatmap) explanation. This indicates that existing explanation failed to indicate for physicians when to rely on AI’s recommendations, and when not to. Otherwise, physicians’ assisted performance would have been higher than either AI or a solo physician. Indeed, by looking into their fine-grained decision change pattern, physicians had initiated both correct and incorrect decision changes that were relatively equivalent in amount, which explains the source of the statistically insignificant difference of accuracies between AI prediction and additional explanation assistance. Prior human-subject studies in the human-computer interaction field observed similar effect of AI explanation in decision support: the AI explanation tends to only increase the chances of human accepting AI’s suggestions, regardless of AI’s correctness [8, 23, 28]. This indicates that additional strategies [16, 34, 30, 20] are needed to carefully craft the design of explainable AI algorithms and interfaces to reduce such overreliance risk on AI [10, 11] and achieve its desired clinical impact.

Qualitative results revealed reasons for the failure of AI explanation to boost physician-AI performance. Physicians had a mixed view of the clinical utility of AI explanation: they saw the heatmap explanation as a useful tool to help them localize important features and easy-to-miss lesions. They also used explanation as a “cross-check” (N3) tool to verify AI decision, calibrate their trust in AI, and ensure the safe use of AI (N1), especially during decision disagreement (Supplemental S1 Section 2). However, when they use AI explanation to verify AI decisions, physicians found that the heatmap explanation only provided limited information on the location of important features, but failed to give explicit reasons, contexts, or descriptions of the highlighted features (Supplemental S1 Section 3.1). This finding echoes similar feedback from pathologists in a user study using heatmap explanations [17]. By providing explanation information to the clinical decision process, our qualitative data analysis further identified that the existing heatmap explanation is missing critical information to construct a clinically relevant explanation [26]: the heatmap explanation neither provides descriptive information on the pathology within image features, nor justifies why and how the highlighted regions lead to the AI’s decisions [19]. Our clinical study revealed the limitations of existing heatmap explanation, and identified clinical usage gaps to improve XAI techniques and achieve their expected clinical utilities.

The ranking of explanation goals revealed a wide range of clinical usage scenarios to use AI explanation in a clinical context. In addition, physicians’ need for explanation showed polarization and varied from person to person, which indicates the use of explanation could be a highly individual choice, and can be presented personally and on demand [22, 24].

## 6 Limitations and future work

Despite the national-scale study, the total number of participants is 35 which is relatively small. Despite our best recruitment effort, we did not get any enrollment of radiologists. This limits further statistical analysis such as multivariate regression analysis to identify variables associated with physicians’ performance improvement. The study is a phase II evaluation using a simulated clinical task on retrospective clinical data. To fully assess the clinical utility of AI and its explanation, future work is needed to conduct a phase III randomized controlled clinical user study on tasks within a real clinical settings, on either retrospective or prospective clinical data.

Future work may improve the existing XAI methods via the following ways: 1) in the technical development phase, XAI developers and researchers should seek more clinical input to understand the clinical reasoning and the physicians’ requirements when incorporating AI assistance in their decision-making process, so that the XAI techniques can potentially improve the joint physician-AI performance; 2) in the clinical deployment phase, additional training or tutorial sessions may be developed to enable clinical users to understand the capability of AI, and incorporate the additional cognitive strategies while interpreting the AI explanation.

## 7 Conclusion

As a fast-advancing technology, AI has the potential to transform neuro-oncological practice and assist physicians in a variety of clinical tasks such as tumor segmentation and disease prediction. To overcome the clinical translational gap of moving AI from bench to bedside, we conducted a phase II evaluation on the clinical utility of AI and its explanation, which is analogous to the phase II clinical trial on the primary efficacy of a new intervention on a small-scale population.

The Canada-wide online survey study recruited 35 neurosurgeons, each of whom read 25 brain MRI from patients with gliomas, and gave their judgment on the glioma grading without and with the assistance of AI’s prediction and explanation. Results showed that, compared to physician performing the task alone, when assisted by AI prediction, physicians’ task performance increased significantly to be equivalent to AI’s performance. But the extra assistance of AI explanation did not bring an additional performance boost. In addition, physicians’ trust in the AI system and willingness to use it increased after viewing AI’s performance metrics compared to the baseline, and such levels did not change after using AI’s predictions and explanations. The study showed the clinical utility of AI assistance in improving physicians’ task performance, and revealed limitations of existing AI explanation techniques.

## Data Availability

Data and code will be made available upon publication.

## Acknowledgements

We thank all the physician participants for their valuable input in the study. We thank Ben Cardoen and Hanene Ben Yedder for help reviewing the manuscript.

## Footnotes

1 http://scipy.org/

2 http://pingouin-stats.org/index.html

