## Supplemental_S1 for "Evaluating the Clinical Utility of Artificial Intelligence Assistance and its Explanation on Glioma Grading Task"

#### Supplementary Material S1: Qualitative Results and Analysis

Weina Jin* Mostafa Fatehi* Ru Guo Ghassan Hamarneh

We present the main findings from the physician user study including the following themes: clinical utility of AI (Section 1), clinical utility of AI explanation (Section 2), and clinical requirements of AI explanation (Section 3). The qualitative data analysis method is presented in Section 4.

We number the participants with N1, N2, ..., or O1, O2, where N indicates neurosurgeon participants in the interview, and O indicates open-ended responses in the online survey. Whenever necessary, we included participants' verbatim quotes despite some minor grammatical errors.

### 1 Clinical utility of AI

#### 1.1 Clinical decision support

The main role of AI mentioned by physicians is to provide clinical decision support, a "second opinion" (N2), or "another level of evidence" (N5). Physicians would not delegate their full clinical tasks to AI at the current stage.

*co-first author

*"I would do my own interpretation, I would see what AI thought, and I would maybe modify mine. but I wouldn't go 100% on AI. I would just use it, there's more information before I made a decision."* (N3)

*"With the original plan the ideal AI program would differentiate that it is a glioma prior to attempting to grade the glioma, that's a very lofty goal. It's a very difficult thing I know as this is. This (current AI) would be useful still in operative planning, if you suspect a glioma, to have this confirmatory step, to say that there's another level of evidence suggesting that it's either high or low grade."* (N5)

##### 1.2 Benefits to junior physicians

The clinical value of AI may be more significant for junior physicians.

*"The whole idea of AI is that a lowly-trained person can do the same job as a highly-trained person. You don't need 10, 20 years of experience, you don't need to have looked at hundreds of thousands of these (MRI cases)."* (N3)

Specifically, for junior physicians, AI can provide the following support:

1. AI provides decision support for time-sensitive cases, and hard cases.

   *"If you want to ask a colleague and no one is available, or if you're trying to do something quickly, and want to make a decision yourself initially, you can just check that decision."* (N2)

   *"This (AI system) would be helpful in asking for a second opinion if someone else is not around, when you're a little bit more uncertain."* (N2)

2. AI improves junior physicians' learning and problem-solving skills: to *"help you reaffirm what you're learning"* (N2).

   *"I think the use cases would probably end up being, junior learners that are trying to learn how to read these scans, that might want to check with someone. Because if you're looking at a scan, you may have an initial thought about what the diagnosis might be, and you might look up that diagnosis, and look up on Radiopedia what the specific features are. This (AI) would be a way to check your thinking instantly."* (N2)

#### 2 Clinical utility of explainable AI

##### 2.1 Color map helps to localize important features and easy-to-miss lesions

All five neurosurgeon interviewees utilized the feature localization information of color maps to 1) inspect important features for an AI decision; and 2) to identify easy-to-miss small focal lesions.

*"I find the color map quite useful for decrypting what the AI is using, its contributory factors."* (N5)

*"If at first pass (of reading MRI) you miss a small FLAIR (pulse sequence) hyper-intensity that doesn't show up with contrast enhancement, that was helpful (for the color map) to help determine."* (N4)

*"The color maps were just pretty, but they didn't explain anything, except for multi-centricity, like if there were multiple tumors, then (the current color map) it's helpful. Here, you can see that there's a second tumor up front, and some people might have missed that. So that way (with the color map), the computer is kind of nudging you, and says, 'Hey look, there's a second lesion.' "* (N3)

*"Take a picture of the (MRI) scan, and it (AI) would then give you another opinion. And then you can either trust what you initially thought, or you know look further into the features that you think are kind of driving your decision, and what the AI is trying to bring your eyes to. That explainability feature was quite cool. Because the other parts of the scan that I had initially missed just based on the fact that I was reviewing the scan pretty quickly, the AI obviously picks it up, where there were tiny little dots of tumor that could have indicated more diffuse spread of the tumor itself, and it can bring your attention to other small foci of tumor that you may have initially missed."* (N2)

*"I think that the AI (and its explanation) helps with a kind of a scanning bias that we have. As humans where we see the large and obvious lesion and we're immediately focused on that. I think that the AI sometimes seem to be better at noticing secondary satellite lesions that definitely changed the diagnostic and probably the prognostic opinion. So a single large lesion can be distracting and keep your attention, whereas smaller secondary lesions I noticed that they were picked up by the color map quite well."* (N5)

#### 2.2 Resolving disagreement

All five neurosurgeons mentioned that explanation is most useful during decision discrepancies between AI and physicians.

*"It's (color map) very helpful when there's discordance between (me and AI). If it's affirming the diagnosis that I've made based on the (MRI) scan, then it's not as relevant. But I would scrutinize it, if it disagreed with me, to determine whether I would change my mind or not. So I think it's not helpful when you're agreeing, but it is helpful to more carefully consider when you disagree."* (N4)

*"I think the use case where it (color map) is very useful is when there's a discordance. So when the algorithm and my opinion disagree, understanding that disagreement is where the color map is very useful."* (N5)

The way doctors resolve decision disagreement is to reassess the explanation together with the inputs by themselves, to see whether the explanation is reasonable according to their clinical prior knowledge. A reasonable explanation that aligns with clinical evidence is more likely to persuade doctors to take AI's suggestion, whereas an explanation that deviates from doctors' own judgment on the contributing features may alert doctors to carefully inspect the original input image and the highlighted areas on the color map, especially to inspect for the easy-to-miss lesions potentially highlighted by the color map. During the inspection, if doctors did not identify any clinically meaningful features from color map localization, doctors will probably negate the validity of the explanation, may not take AI's suggestion despite its high accuracy and potentially correct prediction, and stick to their own judgment for the final decision. The next subsection (§2.3) presents more findings on using explanations to verify AI decisions.

*"That's where the AI's explanation would be valuable (during decision disagreement). Because then I could say, 'Well no, that's a low grade.' And the AI say 'No, it's a GBM.' But if the AI said, 'Well, based on multicentricity and mass effect.' And I'm like, "Oh I missed, oh I didn't see that the AI picked up that there was a second lesion.' Then I would say, 'Oh sorry, I got it wrong. AI got it right."'* (N3)
— A reasonable explanation persuaded doctors to take AI's suggestion.

*"Like the two times we disagreed, I would still stick with mine over AI. But that's after seeing those red (color) maps. Like I said, the red maps were useless. They didn't help at all."* (N3)
— An unreasonable explanation did not persuade doctors to take AI's suggestion.

*"There's only one that I had discordance (with AI prediction), and I think I did change my final judgment depending on looking at the areas that are highlighted. I think I just reevaluated there*

*were areas that they (the color map) had picked up that I had not taken into account, when I scrolled through the (MR) images myself, and the color map would cause me to scrutinize those same areas on the source imaging, so I would look at that color map be, 'okay, what's different about that area that it thinks it's important?' So the highlighted areas of clinical relevance that I missed. I would scrutinize that area a little bit more to see if there was something different about it to sway my decision-making."* (N4)
— Doctor reevaluated his own judgment based on color map localization.

*"I think that it (color map) helped me understand in cases where there was a discordance between what AI was suggesting, why those disagreements might exist. So in cases where there was very little overlap between what I thought were important factors, and what the AI was interpreting based on the color map, I was more confident in disagreeing with the AI. If it had pointed out additional factors that I had not previously noticed, then I was more confident in changing my opinion based on the new points identified. So the disagreement caused me to pause and then go through the images again with the aid of the color map, to see if there was something that I missed. ... So on each individual one, I looked at the components (features) that I thought were important for my diagnosis. I chose my diagnosis, and after that when there was disagreement with the AI, I would go through the (MRI) scan again and the color map, first looking for anything I had missed, and second looking at the components of the color map that the AI felt were important. If the color map aligned with the components that I thought were important, and there was simply a disagreement on grade, based on that, I typically stuck with my own opinion over the opinion of the AI. And if the color map identified factors that I had not seen on my first look at the scan, then I reconsidered, and often changed my opinion."* (N5)
— Doctor's detailed process to identify reasons for decision disagreement based on the alignment of doctor's and AI's important features.

Despite the extra time and effort spent in re-inspecting input image and explanation, a participant still sees the explanation useful to identify doctors' potential errors.

*"Although it does add work, the work that it adds is only in cases where I may have otherwise made an error. So I find it (explanation) very useful as an error checking modality."* (N5)

#### 2.3 Verifying AI decision, and calibrating trust

Since explanation reveals the AI model's decision process, physicians tend to assess the plausibility of explanation based on its agreement with clinical prior knowledge, and use such explanation quality information as a proxy or *"internal validation"* (N1) to judge AI's decision credibility for a given case.

*"This color map didn't work as well as the previous one, and so that's kind of an internal validation to me. I want to see scenarios where it (AI) doesn't work, and I can tell that. So this is reassuring to me that, like they (AI) can make a mistake, and I can call it out, I can determine the mistake. ... Presumably this AI system won't do very well, and then you can show me why it didn't do well, like when you do the explanation, you can say it didn't do well, because it looked at the wrong places."* (N1)

*"It's using relevant points to make that decision, and that in itself would increase my confidence in it. I don't know exactly what it's doing, but I know that it's looking in the same areas and as looking at the patterns of enhancement, and the patterns of high signal within the areas that I'm looking as well, so that intuitively allows me to have some confidence in it. But when you start to see stuff that is in my opinion irrelevant, it makes you question to some extent exactly what it's taking into account in its algorithmic decision-making process."* (N4)

*"It (the color map) might change your confidence level when you're looking at areas which don't appear to have clinical relevance that are being analyzed."* (N4)

Physicians' assessment of color map plausibility directly influences their trust in the specific AI decision, and their overall trust in the AI model in general.

> *"I don't think that it's going to be a binary thing (that color maps can help to judge whether the AI is right or wrong), but I think it (explanation) assists in judging whether or not you can clinically trust the conclusion of the (AI) algorithm."* (N5)

> *"I don't know whether it's correct to use these explanations to kind of triangulate accuracy of the model. So far a lot of the explanations have been factually incorrect, and so the more that I see that there's a discordance between what I think is important and what actually is the tumor, the more I trust my own initial decision of this being a high grade versus a low grade (as predicted by AI).*
>
> *Here you can see the only areas that are red are outside of the tumor. Overall this these explanations have reinforced my initial skepticism about the determination of the model, because even if it used its information and made correct predictions, let's say it was correct for this patient, and I was incorrect. I wouldn't trust it. Because every time every one of these explanations suggests that it's used wrong information to make that decision. It's like prioritizing ventricles over anywhere where the tumor is, even if you (AI) are predicting it correctly, I won't take it that's right.*
>
> *If a system made its prediction based upon these areas (outside the tumor), I would definitely not trust that system, but I would be very reassured that the system is telling me that. ...You show me this (color map), and I see that it's focusing a lot of its decision-making on areas that have nothing to do with the tumor. That decreases the trust in that specific model (AI decision on this specific case), but it increases the overall trust in AI. So I'm less likely to use this model, but I'm more likely to use a model that does a better job than this, because I am reassured that when I see that better model, that I will be able to have access to that back-end explanation. So you're not asking me to believe in the system without evidence."* (N1)

> *"There were areas which were clearly had nothing to do with the decision making which were clearly outliers, like it would often pick up interhemispheric frontal and show me that as part of the color map, or temporal poles and stuff like that where you might get artifact, and that has nothing to do with the decision making. So they're kind of areas which seem like red herrings kind of irrelevant, and I don't know why it would be using some of that in its decision-making process. So the only thing is that makes me feel a little less confident in this machine learning ability, because it's showing areas that don't seem relevant to making the decision presented to me."* (N4)

Explanation was also regarded by clinicians as a "debugging" tool for physicians to verify AI decisions and cross-check whether AI is malfunctioning. This is an important clinical utility of explanation to *"ensure safety"* (N1) in AI-assisted *"life-and-death clinical decisions"* (N3).

> *"I think it's gonna be a big leap one day, if you just press a button, and the AI says it's GBM. Well, clinicians are gonna want an explanation. We're not like, because what if there's a malfunction in the computer? We like to cross-check. And if the AI said, 'based on the contrast enhancement, based on the T2 in this case showed that edema, then that is why the AI thinks it's a high likelihood.' Then I could look at the pictures, I see what the AI was thinking, 'Okay, good, I agree.' So that's the ideal expect."* (N3)

> *"If the model keeps making the same mistake for a subset of patients, it's then important for you to be able to tell me how it (AI) made that mistake, and that's a utility of explanation."* (N1)

#### 2.4 Making medical discoveries

One physician mentioned that the explanation can be used to make new medical discoveries by identifying new features that are different from humans' pattern recognition, providing the AI has a good performance, and the explanation truthfully expresses the AI model's decision process.

*"The other fascinating thing is that, maybe there is a lot of information encoded in a different dimension than what we look at. Because as humans we use our own pattern recognition, and we've gotten very good at differentiating contrast enhancement and necrosis. But maybe there's more information to be gathered from the AI perspective. ... So helping you to identify patterns and features that I would have previously not thought are important. So for example, if you tell me an AI machine that every time it picks up a lot of weird things on FLAIR (pulse sequence), and then has the best accuracy, and it makes you think maybe there is something important on the FLAIR that we haven't been paying attention to. So that's the added value of explanation."* (N1)

### 3 Clinical requirements of explainable AI

#### 3.1 Limitations of existing color map explanation

We analyze the limitations of the existing color map explanation by corresponding the information provided by the form of color map to the clinical image interpretation process. In general, doctors' image interpretation consists of two steps: 1) First, they perform pattern recognition and extract key features. This includes recognizing or localizing key features, and identifying their pathology. And then 2) they perform medical reasoning and construct multiple diagnostic hypotheses (differential diagnosis) based on the image feature evidence.

*"What (explanation) we get currently, when a radiologist read it, they point out the significant features, and then they integrate those knowledge, and say, to my best guess, this is a GBM. And I have the same expectations of AI (explanation)."* (N3)

A complete explanation may correspond to the above clinical image interpretation process. A clinically relevant explanation at least corresponds to the aforementioned step 1), i.e., identifying important features and describing their pathology. The existing form of color map explanation, however, only localizes important features, but lacks the description of the pathology of important features. This fatal drawback of color map explanation made all physician participants confused, and they requested an explanation for the meaning of the highlighted regions in the color map.

*"What does that (color map region) mean? Like hey, which part of my car gets my car moving? It should say press the accelerator. But yours would just show a dashboard of the car, and show that the accelerator had a little bit of red on it, this button had some red, that button had some red, but it's not an explanation. A picture is never an explanation. Like more red indicates the region is more important, what about that region? Like go to an example, and you'll see, what about the red areas under MRI T1CE (pulse sequence)? Was it central necrosis? But it couldn't be the central necrosis, because there's more central necrosis in the temporal lobe, and that area didn't get highlighted. So anyway, I don't know, it's just confusing.*

*These color maps were totally useless without text, without any context or explanation, like those details. The color maps were just pretty, but they didn't explain anything."* (N3)

*"Though the color map is drawing your eyes to many different spots, but I feel like I didn't understand why my eyes were being driven to those spots, like why were these very specific components important? And I think that's where all my confusion was."* (N2)

In addition, since the color map cannot explain the reasons for localizing clinically irrelevant features, physicians preferred to see an explanation that aligns with their prior knowledge, simply to avoid confusion during the interpretation of clinically irrelevant features.

*"My priority would be to have the AI show and explain the features inside the tumor that are important. Because I'm not sure what it's capturing outside the tumor that is important to the (AI) system actually."* (N2)

*"There are quite a few areas that tend to 'light up' on the prediction map that are false positives. And it would be good to know where/why these false positive areas are interpreted as such."* (O1)

*"Although this appears to do a reasonable job at predicting disease grade, it does not appear to diagnose the presence of a lesion versus normal, nor to have the reliability required to forego a biopsy."* (O2)

#### 3.2 Desirable explanation

Physicians described the ideal explanation could be some simple linear or rule models, with clinically relevant features as the variables.

*"I know it's an AI model, so it's not like a regression model. But it would be helpful to discuss what variables the model is using. ... So maybe an explanation beginning with factors: the enhancement pattern, the midline shift, the amount of edema, more like radiologists language, that's what clinicians would use to guess if it's GBM or not."* (N3)
— Use clinically relevant features as variables for the explanation model

*"If this was like a logistic regression model, you could say, well based on the fact that it's enhancement: yes/no? yes, multicentricity: yes/no? yes, and then mass effect: yes/no, yes, edema: yes. You got four yeses, so your chances of having this be GBM is 94%. So like you use old-school statistics to explain."* (N3)
— Use a linear model as explanation

*"I want to be able to work with the (AI) model to go down like a flow chart to, I understand that's not how necessarily machine learning works, but if you're trying to explain, or show the relevant areas if I had some degree of insight into how the model was trained, or what the relative importance of each of these color maps was in making its decision, then it might help me build confidence with it."* (N4)
— Use rule-based explanation to dissect the decision

In addition, compared with a global explanation that explains the AI model's behavior in general, doctors have more demand for a local explanation for a specific case.

*"I expected the explanations to be different for every patient, every scan (the explanation) should be different."* (N3)

#### 3.3 Making AI transparent by providing information on performance, training dataset, and decision confidence

Besides the color map explanation, some physicians mentioned that other information is required to make the AI model decision process transparent, and help build trust in the AI model and its suggestions. The requested information includes model performance, information on training and validation dataset (such as "*distribution of the patient's demographics, distribution of lesions diagnosed*" (N5)), and decision uncertainty overall and at the MRI pulse sequence level for a given case.

*"The trust in the AI model comes from performance ultimately. Color maps are secondary to the performance. So if you tell me system A has a 99% accuracy, and system B has a 22% accuracy. I don't care how it figured that out as long as it (has high accuracy). Like I said, clinicians are numbers people. If you tell me over a thousand patients this did really well, and this one didn't do really well, I'm okay with that black box designation. If you tell me that for this study or this system is 85 (percent accuracy), and you're asking me how likely I am to trust it, then I think about, 'okay, let's see how it made that decision.' So explain to me how you did that, and that's the value that you're adding with the explainable AI."* (N1)

*"I'm not sure what the relative importance of each one of those color maps is in making its decision. So if it had like, as it could express to me it's confidence for each of those images of it being a high-grade glioma, and then its overall confidence based upon those individual color maps. Because all of this is based on a threshold model of confidence. So if I have an idea about what that threshold is, saying there's a 92% chance this is a high-grade glioma, and these are the important areas, or these are the relevant areas that make that confidence. Because for me, I don't know how much that color map relates to the confidence overall in its prediction. ... I think as someone who reads (MRI) scans and gives diagnosis, we are very happy to deal with probabilities. I imagine there's just a threshold that has been reached that you're using that as a confidence for the model. So if I knew what that threshold was, and how close it (the prediction) was to that threshold, then it would help me at least understand a little bit more about the model, and having that transparency in the models. Because if we were to do this without AI, and if you're sitting down with a few radiologists in a room, they would all tell you what they think it is, and explain why it is that, and then you could question them to say, 'Okay, what is your likelihood? Or what's in your differential diagnosis? What are your relative probabilities of each of these things in your differential diagnosis?' "* (N4)

*"I know that a data set that's not trained properly does not come with good answers, so to calibrate my trust, I need to know that the data set that was used is relevant to the data that I'm looking at to making sure that I'm inputting the same, that my scan is no different than (the training dataset). In a clinical trial, you need to know your patient that is in front of you is of similar demographic and condition as the people who are in a clinical trial to determine how applicable it is. It's the same thing with any AI model is that, you need to know that the AI model is representative of what you're looking at at the same time. Because without that, it's not relevant in you (task), and you can't trust it."* (N4)

### 4 Qualitative Data Analysis Method

We analyzed the qualitative data including the interview transcript and open-ended questions in the survey using an inductive thematic analysis approach [1]. A total of 180 minutes of interviews were recorded and transcribed. We performed open coding on the qualitative data. A total of 85 codes were generated. We then performed an axial coding and affinity diagram process to discover the hierarchical structure among the emerging concepts. The first author who has expertise in clinical medicine and human-computer interaction research conducted the interview and qualitative data analysis process.
