## Supplemental_S2 for "Evaluating the Clinical Utility of Artificial Intelligence Assistance and its Explanation on Glioma Grading Task"

#### Supplemental S2: AI Model and Explanation, Additional Quantitative Results

Weina Jin* Mostafa Fatehi* Ru Guo Ghassan Hamarneh

*co-first author

### 1 Training AI Model and Generating Explanation

#### 1.1 AI model and algorithmic evaluation on glioma grading task

We trained an AI model using the BraTS 2020 dataset to grade glioma MRIs. The AI model receives an MRI input and outputs a glioma grade of either a grade II/III glioma or a glioblastoma (GBM). The MRI input has the size of $4 \times 240 \times 240 \times 155$, which are the number of pulse sequences, height, width, and depth, respectively. The model architecture is a VGG-like [13] three-dimensional (3D) convolutional neural network (CNN), with six 3D CNN layer connected to two fully connected layers. During model training, to overcome the class imbalance issue, we used a weighted sampler to sample grade II/III glioma or GBM class weighted by their inverse sample count. We used the cross-entropy loss function, and trained the model with an Adam optimizer, a learning rate of 0.0005, a batch size of 4, and 32 epochs.

We stratified split the BraTS dataset into 65% training (239 cases), 15% validation (56 cases), and 20% (74 cases) hold-out test set by keeping the same grade II/III glioma: GBM ratio in each set. There were no patient's ID overlapping among the three datasets. We used the training data to train AI models, the validation to select the hyperparameters and best-performing model, and the hold-out test data to report AI model performance. The training, validation, and test accuracies of the AI model are 80.28%, 92.86%, and 90.54%, respectively. The fine-grained model performance metrics are in Fig. 1, which was also shown to participants in the clinical study.

We used PyTorch[1] and MONAI API[2] for model training, and Captum[3] to generate post-hoc color maps. To train the models and generate color maps, we used a computer with 1 GTX Quadro 24 GB GPU and 8 CPU cores, and a SLURM[4] based high performance computing cluster with jobs configured to use no more than a minimum of 1 GPU, and 8 cores CPU each with 128 RAM.

[1]http://pytorch.org
[2]http://monai.io
[3]http://captum.ai
[4]https://slurm.schedmd.com/overview.html

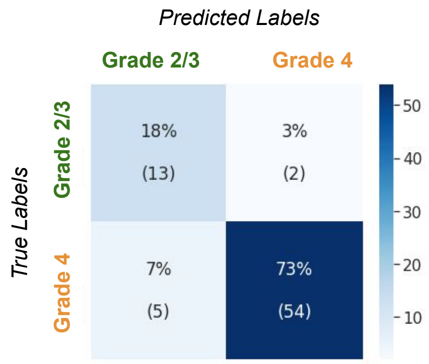

| | Sensitivity | Specificity | Positive Predictive Value | F1 score | Number of class in the test set |
|---|---|---|---|---|---|
| **Grade 2 or 3 glioma** | 0.87 | 0.92 | 0.72 | 0.79 | *15* |
| **Grade 4 glioblastoma** | 0.92 | 0.87 | 0.96 | 0.94 | *59* |

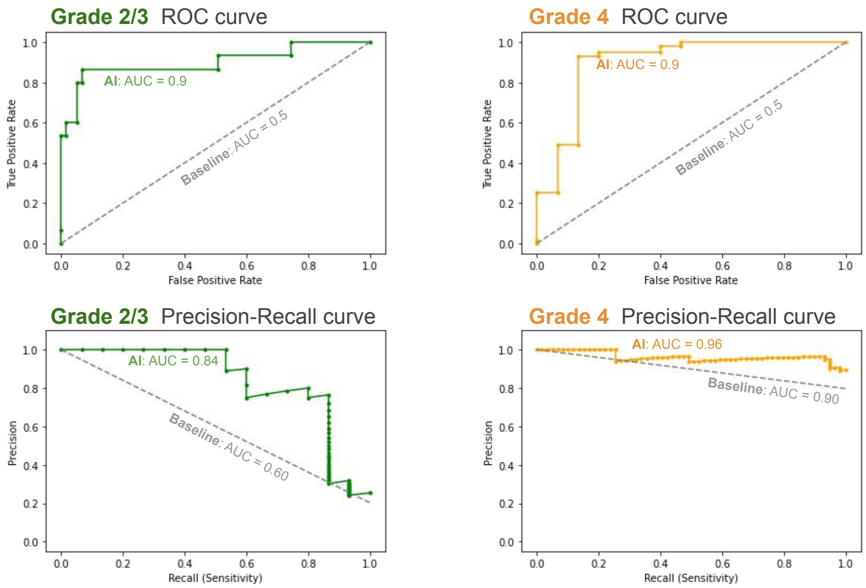

Figure 1: AI model performance metrics. A simplified version was also shown in the clinical study.

#### 1.2 Generating and selecting the optimal AI explanation

The AI model we trained to grade glioma is a black-box CNN model. To explain the model decisions to physicians, we applied post-hoc XAI algorithms that act as a surrogate model to approximate the black-box AI model by probing the model parameters and/or input-output pairs. We aimed to select the optimal XAI algorithm to use in the clinical study from a candidate list of 16 post-hoc XAI algorithms: Gradient [12], Guided BackProp [15], Deconvolution [17], SmoothGrad [14], GradCAM [9], Guided GradCAM [9], Input×Gradient [11], DeepLift [10], Integrated Gradients [16], Gradient Shap [7], Occlusion [17, 18], Feature Ablation [1], Feature Permutation [3], Lime [8], Shapley Value Sampling [2], and Kernel Shap [7]. These algorithms belongs to the feature attribution method. They generate a feature attribution map or color map overlaid on the input image, to explain the important image regions for model prediction. The selection criterion is to choose the most truthful XAI algorithm to the AI model decision process [5, 4]. Following the cumulative feature removal method to evaluate XAI truthfulness in Jin et al. [5, 6], we conducted a computational evaluation to calculate the $\Delta$AUPC score from the cumulative feature removal method of the 16 XAI algorithms on the test set. The cumulative feature removal method iteratively removes the input from the most to the least important features according to the color map explanation, and plots the relationship between gradual feature ablation and model accuracy. The evaluation metric $\Delta$AUPC is to quantify the degree of performance deterioration by calculating the difference of area under the perturbation curve (AUPC) between an XAI algorithm $\mathcal{H}$ and its random feature removal baseline $\mathcal{H}_b$. $\Delta$AUPC is in the range of $[-1, 1]$, with a high $\Delta$AUPC indicating a more truthful explanation. SmoothGrad had the highest $\Delta$AUPC score of 0.33, thus it was the most truthful XAI method among the 16 XAI algorithms. We chose to use SmoothGrad as the optimal XAI method to generate AI model explanations in the clinical study. Two SmoothGrad explanation color maps used in the clinical study are visualized in Fig. 2. The result of the cumulative feature removal experiment is shown in Fig. 3.

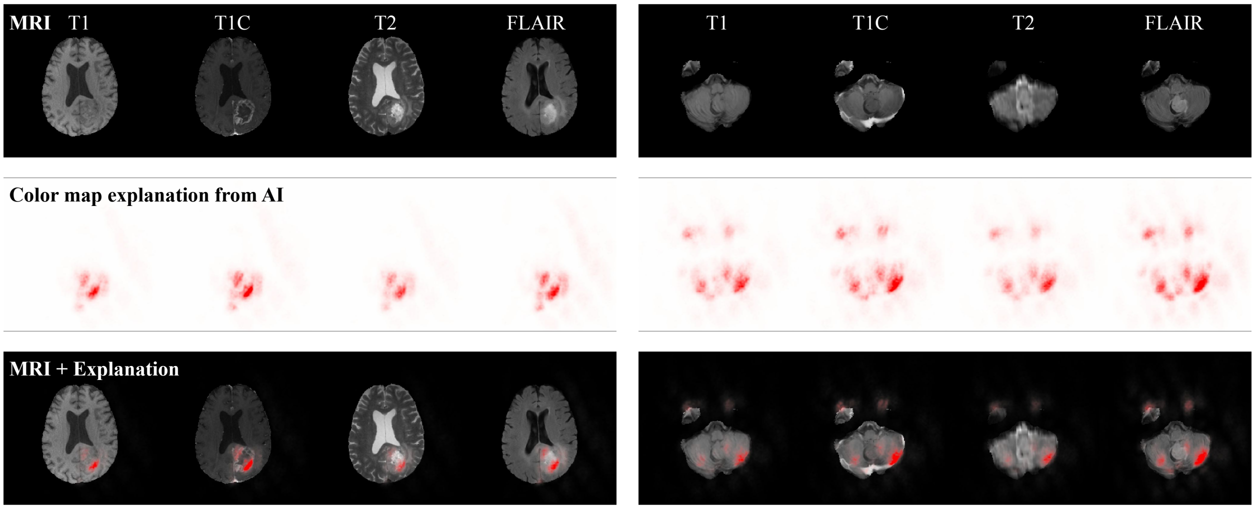

Figure 2: Visualization of two MRIs and their corresponding color map explanations used in the study. We include two cases that have the highest (left, GBM, data id: BraTS20_Training_056) and lowest (right, grade II/III glioma, data id: BraTS20_Training_325) average rating on explanation quality. AI predicts correctly for both MRI and the color maps show AI explanation on the prediction. Each column is MRI pulse sequence of T1, T1C, T2, and FLAIR. Row 1 is MRI image, Row 2 is color map, and Row 3 is color map overlaid on the MRI. The original MRIs and color maps are both 3D images shown as one 2D image in axial view.

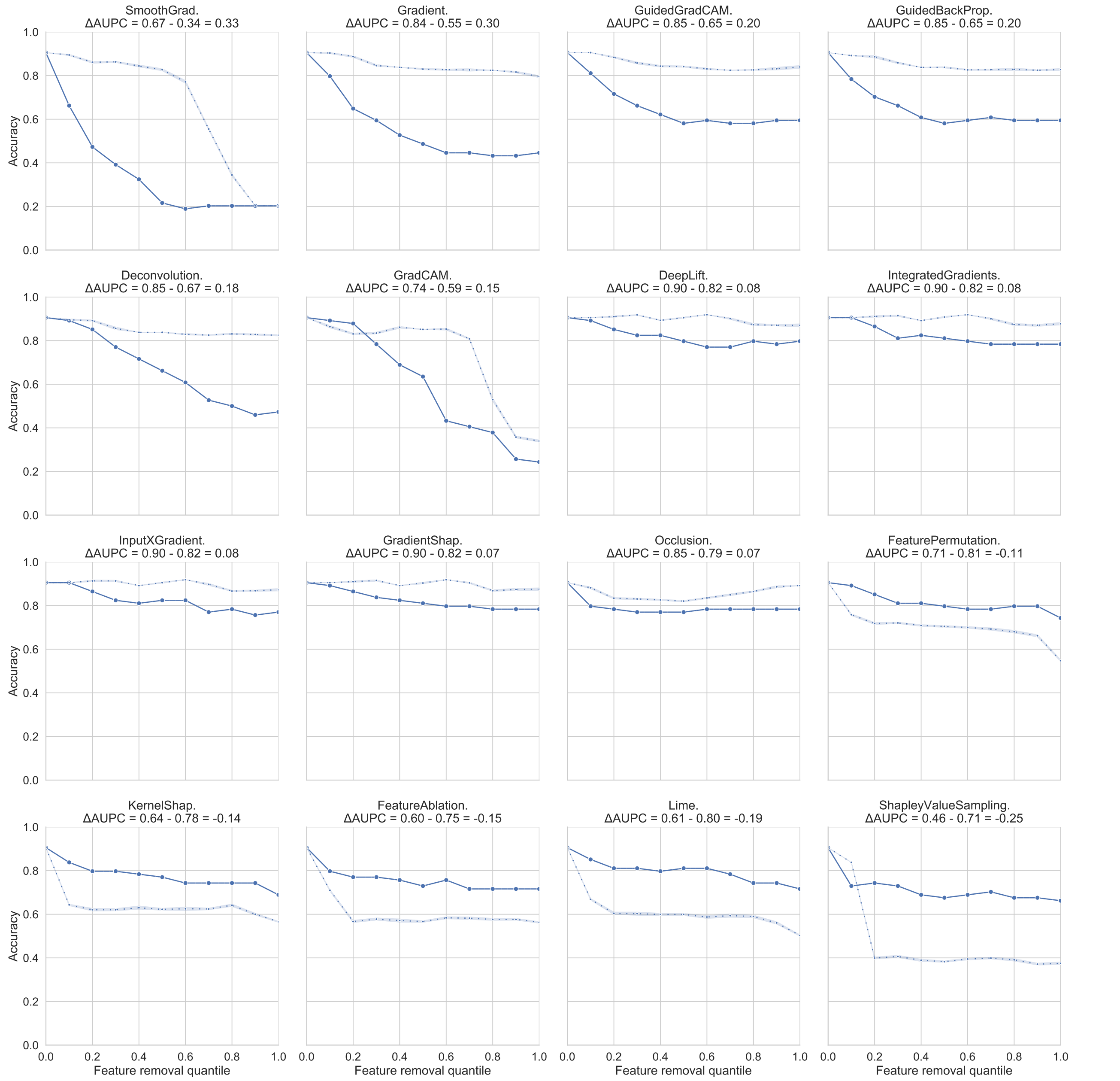

Figure 3: **Feature perturbation curve of cumulative feature removal experiment on the glioma task.** Each plot shows the feature perturbation curve of an XAI method ($\mathcal{H}$, solid line) and its random baseline ($\mathcal{H}_b$, dashed line). A bigger gap between the two curves indicates a higher $\Delta$AUPC, thus a better performance on explanation truthfulness. Plots were arranged according to their $\Delta$AUPC value ($\Delta\text{AUPC}(\mathcal{H}) = \text{AUPC}(\mathcal{H}_b) - \text{AUPC}(\mathcal{H})$, with numbers rounded to two decimal places) as indicated in the plot subtitle.

### 2 Additional Quantitative Results

#### 2.1 Participants

| DR ID | DR | DR + AI | DR + XAI | MRI num | Need XAI (%) | XAI Qual. | Position | Yr | Age | Gender | AI Familiarity | AI Attitude |
|---|---|---|---|---|---|---|---|---|---|---|---|---|
| 01 | 80.00 | **84.00** | 80.00 | 25 | 16.67 | $5.68 \pm 3.40$ | Resident | 1 | 31 | Female | use AI in work/life | Interested |
| 02 | 80.00 | **92.00** | 92.00 | 25 | 12.50 | $9.00 \pm 1.06$ | Resident | 5 | 30 | Female | hear of AI | Interested |
| 03 | 84.00 | 80.00 | 80.00 | 25 | 96.00 | $6.21 \pm 2.24$ | Resident | 4 | 32 | Male | hear of AI | Neutral |
| 04 | 82.35 | **94.12** | 94.12 | 17 | 17.65 | $5.62 \pm 1.76$ | Resident | | | | | |
| 05 | 88.00 | 88.00 | **92.00** | 25 | 68.00 | $8.29 \pm 1.51$ | Attending | 5 | 44 | Male | hear of AI | Interested |
| 06 | 76.00 | **84.00** | **88.00** | 25 | 12.00 | $5.25 \pm 1.53$ | Attending | 4 | 37 | Male | hear of AI | Skeptical |
| 07 | 100.00 | 100.00 | 100.00 | 2 | 0.00 | $7.50 \pm 0.50$ | Attending | | | | | |
| 08 | 68.00 | 68.00 | **72.00** | 25 | 33.33 | $5.24 \pm 1.24$ | Attending | 13 | 47 | Male | hear of AI | Skeptical |
| 09 | 80.00 | 80.00 | 80.00 | 25 | 20.00 | $1.12 \pm 0.43$ | Resident | 8 | 30 | Female | never hear of AI | Not interested |
| 10 | 84.00 | **96.00** | 92.00 | 25 | 28.00 | $5.04 \pm 2.07$ | Resident | 7 | 30 | Female | hear of AI | Skeptical |
| 11 | 88.00 | 88.00 | 88.00 | 25 | 0.00 | $5.68 \pm 2.29$ | Resident | 9 | 35 | Male | can write AI code | Excited |
| 12 | 100.00 | 100.00 | 100.00 | 3 | 66.67 | $2.00 \pm 0.00$ | Fellow | | | | | |
| 13 | 60.00 | **76.00** | 76.00 | 25 | 0.00 | $7.72 \pm 2.46$ | Resident | 4 | 31 | Female | hear of AI | Neutral |
| 14 | 88.00 | 84.00 | **88.00** | 25 | 25.00 | $5.71 \pm 2.07$ | Attending | 30 | 62 | Male | use AI in work/life | Excited |
| 15 | 66.67 | **100.00** | 100.00 | 2 | 100.00 | $5.00 \pm 0.00$ | Attending | | | | | |
| 16 | 84.00 | 84.00 | 84.00 | 25 | 20.00 | $6.40 \pm 2.24$ | Resident | 4 | 29 | Male | can program, but not write AI code | Interested, Excited |
| 17 | 87.50 | 87.50 | **100.00** | 8 | 25.00 | $7.25 \pm 2.17$ | Resident | | | | | |
| 18 | 88.00 | **96.00** | 88.00 | 25 | 48.00 | $8.28 \pm 1.43$ | Resident | 4 | 28 | Female | hear of AI | Interested |
| 19 | 80.00 | **92.00** | 92.00 | 25 | 100.00 | $8.68 \pm 2.19$ | Resident | 1 | 27 | Female | use AI in work/life | Interested |
| 20 | 80.00 | 80.00 | **84.00** | 25 | 88.00 | $5.68 \pm 2.38$ | Resident | 4 | 31 | Male | use AI in work/life | Excited |
| 21 | 88.00 | 88.00 | 88.00 | 25 | 24.00 | $7.00 \pm 2.83$ | Resident | 4 | 29 | Male | can program, but not write AI code | Neutral |
| 22 | 88.00 | 88.00 | 88.00 | 25 | 0.00 | $1.48 \pm 0.77$ | Attending | | | | hear of AI | |
| 23 | 84.00 | 84.00 | 84.00 | 25 | 16.00 | $1.04 \pm 0.77$ | Attending | 23 | | | use AI in work/life | Not interested |
| 24 | 92.00 | 92.00 | 92.00 | 25 | 24.00 | $6.48 \pm 1.70$ | Resident | 0 | 27 | Male | can write AI code | Interested, Excited |
| 25 | 66.67 | **100.00** | 100.00 | 2 | 50.00 | $9.50 \pm 0.50$ | Resident | | | | | |
| 26 | 80.00 | **84.00** | 84.00 | 25 | 20.00 | $6.60 \pm 2.61$ | Attending | 6 | 38 | Male | | Excited |
| 27 | 84.00 | 84.00 | 84.00 | 25 | 13.04 | $6.56 \pm 2.10$ | Attending | 5 | 44 | Male | hear of AI | Interested, Excited |
| 28 | 68.00 | **80.00** | **88.00** | 25 | 16.00 | $5.92 \pm 1.83$ | Resident | | | | can program, but not write AI code | |
| 29 | 88.00 | **92.00** | 92.00 | 25 | 8.70 | $7.83 \pm 1.03$ | Resident | 3 | 31 | Male | hear of AI | Interested |
| 30 | 88.00 | **92.00** | **96.00** | 25 | 96.00 | $6.44 \pm 1.50$ | Resident | 7 | 30 | Male | use AI in work/life | Interested |
| 31 | 80.00 | **88.00** | 88.00 | 25 | 96.00 | $7.80 \pm 2.94$ | Fellow | 7 | 33 | Male | can program, but not write AI code | Interested, Excited |
| 32 | 80.00 | 80.00 | 80.00 | 25 | 12.00 | $8.42 \pm 2.52$ | Attending | 11 | 40 | Male | hear of AI | Interested |
| 33 | 80.00 | **84.00** | 84.00 | 25 | 0.00 | $6.20 \pm 1.77$ | Resident | 4 | 31 | Male | can program, but not write AI code | Interested |
| 34 | 88.00 | **92.00** | 92.00 | 25 | 16.00 | $2.56 \pm 0.57$ | Attending | 15 | 48 | Male | hear of AI | Skeptical |
| 35 | 88.00 | 88.00 | 88.00 | 25 | 100.00 | $10.00 \pm 0.00$ | Resident | 4 | 28 | Male | can program, but not write AI code | Interested |

Table 1: Participants' demographics and their accuracies (%) in three conditions: 1) DR: without AI assistance, 2) DR+AI: with the assistance of AI prediction, and 3) DR+XAI: with the assistance of both AI prediction and explanation. We mark the accuracy in bold in DR+AI column if it is higher than DR; similarly, we mark the accuracy in bold in DR+XAI column if it is higher than DR+AI. MRI num column indicate the number of MRIs participant interpreted in the survey. Need XAI is the percentage of participants need to check AI explanation for the MRI case. XAI Qual. is the mean±std rating from participants on the explanation quality for each color map explanation on a [0, 10] scale. Participants' demographics, including their position, years of experience in neurosurgery (Yr), familiarity with AI, and attitude toward AI are also listed.

| Data ID | GT | AI Pred. | DR | DR + AI | DR + XAI | DR num | Need XAI (%) | XAI Qual. | MRI link |
|---|---|---|---|---|---|---|---|---|---|
| BraTS20_Training_221 | 1 | 1 | 100.00 | 100.00 | 100.00 | 35 | 50.00 | $7.23 \pm 2.75$ | vimeo.com/558775183 |
| BraTS20_Training_208 | 1 | 1 | 100.00 | 100.00 | 100.00 | 30 | 26.67 | $6.79 \pm 2.58$ | vimeo.com/558775334 |
| BraTS20_Training_116 | 1 | 1 | 100.00 | 100.00 | 100.00 | 30 | 16.67 | $6.66 \pm 2.69$ | vimeo.com/558785220 |
| BraTS20_Training_114 | 1 | 1 | 93.55 | 93.55 | 93.55 | 31 | 35.48 | $6.55 \pm 2.47$ | vimeo.com/558795281 |
| BraTS20_Training_112 | 1 | 1 | 93.55 | **100.00** | 100.00 | 31 | 25.81 | $6.93 \pm 2.72$ | vimeo.com/558795007 |
| BraTS20_Training_099 | 1 | 1 | 19.35 | **35.48** | **41.94** | 31 | 76.67 | $3.90 \pm 2.68$ | vimeo.com/558764148 |
| BraTS20_Training_094 | 1 | 1 | 80.65 | **93.55** | 90.32 | 31 | 30.00 | $6.39 \pm 2.87$ | vimeo.com/558764221 |
| BraTS20_Training_093 | 1 | 1 | 100.00 | 100.00 | 100.00 | 30 | 24.14 | $7.13 \pm 2.39$ | vimeo.com/558768608 |
| BraTS20_Training_289 | 0 | 0 | 86.21 | **93.10** | **96.55** | 29 | 37.93 | $5.50 \pm 2.80$ | vimeo.com/558897027 |
| BraTS20_Training_076 | 1 | 1 | 96.77 | **100.00** | 100.00 | 31 | 26.67 | $6.52 \pm 2.89$ | vimeo.com/558768466 |
| BraTS20_Training_075 | 1 | 1 | 96.67 | **100.00** | 96.67 | 30 | 26.67 | $6.61 \pm 2.79$ | vimeo.com/558768604 |
| BraTS20_Training_070 | 1 | 1 | 80.00 | **90.00** | **96.67** | 30 | 30.00 | $6.90 \pm 2.59$ | vimeo.com/546342295 |
| BraTS20_Training_325 | 0 | 0 | 87.10 | **93.33** | 93.33 | 30 | 37.93 | $3.13 \pm 2.85$ | vimeo.com/558895720 |
| BraTS20_Training_064 | 1 | 1 | 100.00 | 100.00 | 100.00 | 30 | 20.00 | $6.93 \pm 2.50$ | vimeo.com/558762879 |
| BraTS20_Training_063 | 1 | 1 | 48.39 | **64.52** | **67.74** | 31 | 50.00 | $6.10 \pm 2.72$ | vimeo.com/558762829 |
| BraTS20_Training_060 | 1 | 1 | 89.66 | **96.55** | 96.55 | 29 | 37.93 | $6.38 \pm 2.59$ | vimeo.com/558758233 |
| BraTS20_Training_056 | 1 | 1 | 96.67 | **100.00** | 100.00 | 30 | 23.33 | $7.27 \pm 2.64$ | vimeo.com/558758468 |
| BraTS20_Training_053 | 1 | 1 | 100.00 | 100.00 | 100.00 | 30 | 20.69 | $7.20 \pm 2.50$ | vimeo.com/558758697 |
| BraTS20_Training_270 | 0 | 1 | 0.00 | 0.00 | 0.00 | 29 | 17.24 | $6.65 \pm 2.64$ | vimeo.com/558784455 |
| BraTS20_Training_277 | 0 | 0 | 41.94 | **61.29** | 61.29 | 31 | 43.33 | $6.13 \pm 2.58$ | vimeo.com/546658075 |
| BraTS20_Training_269 | 0 | 0 | 96.67 | **100.00** | 100.00 | 30 | 30.00 | $5.63 \pm 3.04$ | vimeo.com/558775144 |
| BraTS20_Training_264 | 0 | 0 | 100.00 | 100.00 | 100.00 | 29 | 24.14 | $5.29 \pm 2.76$ | vimeo.com/558775501 |
| BraTS20_Training_280 | 0 | 0 | 83.87 | **90.32** | 90.32 | 31 | 34.48 | $5.74 \pm 2.88$ | vimeo.com/558774958 |
| BraTS20_Training_171 | 1 | 0 | 83.33 | 70.00 | **76.67** | 30 | 65.52 | $5.07 \pm 2.82$ | vimeo.com/558769028 |
| BraTS20_Training_212 | 1 | 0 | 83.33 | 70.00 | 66.67 | 30 | 58.62 | $4.13 \pm 2.88$ | vimeo.com/558786569 |

Table 2: We list individual MRI, their ground-truth label (the column GT) of grade II/III glioma (label 0) or GBM (label 1), their predicted label from AI (AI Pred.), and the participants' accuracies (%) in three conditions: 1) DR: without AI assistance, 2) DR+AI: with the assistance of AI prediction, and 3) DR+XAI: with the assistance of both AI prediction and explanation. We mark the accuracy in bold in DR+AI column if it is higher than DR; similarly, we mark the accuracy in bold in DR+XAI column if it is higher than DR+AI. DR num is the number of collected responses from participants. Need XAI is the percentage of participants need to check AI explanation for the MRI case. XAI Qual. is the mean±std rating from participants on the explanation quality for each color map explanation on a [0, 10] scale. The MRI and its color map explanation can be viewed by following the video links in the MRI link column.

#### 2.2 Physicians' task performance with and without the assistance of AI and its explanation

Since the task of glioma grading is concrened about the correctness of prediction, in the manuscript, we report the results using the performance metric of accuracy. As a supplemental, we also report results using other performance metrics, including F1, Matthews correlation coefficient (MCC), sensitivity, specificity, and positive predictive value (also called precision) in Table 3. Statistical test show similar trends as the result reported using accuracy: using Friedman tests, there are statistically significant differences in task performance measured by a particular metrics among the three conditions. Post-hoc analysis using Wilcoxon signed-rank tests with Bonferroni correction show that, DR+AI condition had a statistically higher performance compared to the DR condition; similarly, the DR+XAI condition had a statistically higher performance compared to the DR condition. However, the performances between DR+AI and DR+XAI conditions does not show statistically significant difference.

| Metric | AI | DR | DR+AI | DR+XAI | $p$-value: Friedman | $p$-value: DR vs. DR+AI | $p$-value: DR vs. DR+XAI | $p$-value: DR+AI vs. DR+XAI |
|---|---|---|---|---|---|---|---|---|
| **Acc.** | 0.8800 | 0.8249 ± 0.0869 | 0.8770 ± 0.0733 | 0.8852 ± 0.0702 | 7.755e-06 | 0.001543 | 0.0004037 | 0.3454 |
| **F1 grade II/III** | 0.8000 | 0.6424 ± 0.2257 | 0.6997 ± 0.2356 | 0.7116 ± 0.2390 | 4.341e-05 | 0.002134 | 0.0006973 | 0.5456 |
| **F1 GBM** | 0.9143 | 0.8777 ± 0.0644 | 0.9126 ± 0.0559 | 0.9187 ± 0.0535 | 7.755e-06 | 0.002895 | 0.0008773 | 0.6383 |
| **MCC** | 0.7181 | 0.5448 ± 0.2274 | 0.6242 ± 0.2340 | 0.6415 ± 0.2365 | 4.341e-05 | 0.002134 | 0.0006985 | 0.5456 |
| **Sen. grade II/III/Spec. GBM** | 0.8571 | 0.6694 ± 0.2444 | 0.7184 ± 0.2510 | 0.7224 ± 0.2521 | 0.0001117 | 0.0178 | 0.01081 | 0.9519 |
| **Sen. GBM/Spec. grade II/III** | 0.8889 | 0.8800 ± 0.0961 | 0.9060 ± 0.0870 | 0.9156 ± 0.0834 | 0.001765 | 0.05405 | 0.008309 | 0.7735 |
| **PPV grade II/III** | 0.7500 | 0.6412 ± 0.2502 | 0.7041 ± 0.2608 | 0.7226 ± 0.2635 | 4.341e-05 | 0.006771 | 0.001361 | 0.7418 |
| **PPV GBM** | 0.9412 | 0.8845 ± 0.0751 | 0.9239 ± 0.0472 | 0.9263 ± 0.0455 | 2.901e-05 | 0.001361 | 0.0006973 | 0.6061 |

Table 3: Physicians' task performance with physician alone (DR), with the assistance of AI prediction (DR+AI), and with the assistance of AI prediction and its explanation (DR+XAI), using multiple performance metrics. We also list AI's performance using these performance metrics (AI), the $p$-value of the Friedman test on the performance differences among the three conditions (DR, DR+AI, DR+XAI), and the $p$-values using Wilcoxon signed-rank tests with Bonferroni correction on the pairwise conditions.

In addition to the task performance analysis on the participants' data as a whole, we performed subgroup analysis by dividing participants into two groups according to their clinical positions and experience: 1) attending physicians, and 2) resident and fellow physicians. The descriptive statistics of the task performance accuracies for each subgroup are shown in Table 4 and 5.

| Condition | N | M±SD | Min | 25% Q | Mdn | 75% Q | Max |
|---|---|---|---|---|---|---|---|
| **DR** | 12 | 82.56 ± 9.28 | 66.67 | 79.00 | 84.00 | 88.00 | 100.00 |
| **DR+AI** | 12 | 86.33 ± 8.61 | 68.00 | 84.00 | 84.00 | 89.00 | 100.00 |
| **DR+XAI** | 12 | 87.67 ± 7.90 | 72.00 | 84.00 | 88.00 | 92.00 | 100.00 |

Table 4: Descriptive statistics for attending physicians' task performance accuracy (%). N - number of participants, M - mean, SD - standard deviation, Q - quantile, Mdn - median.

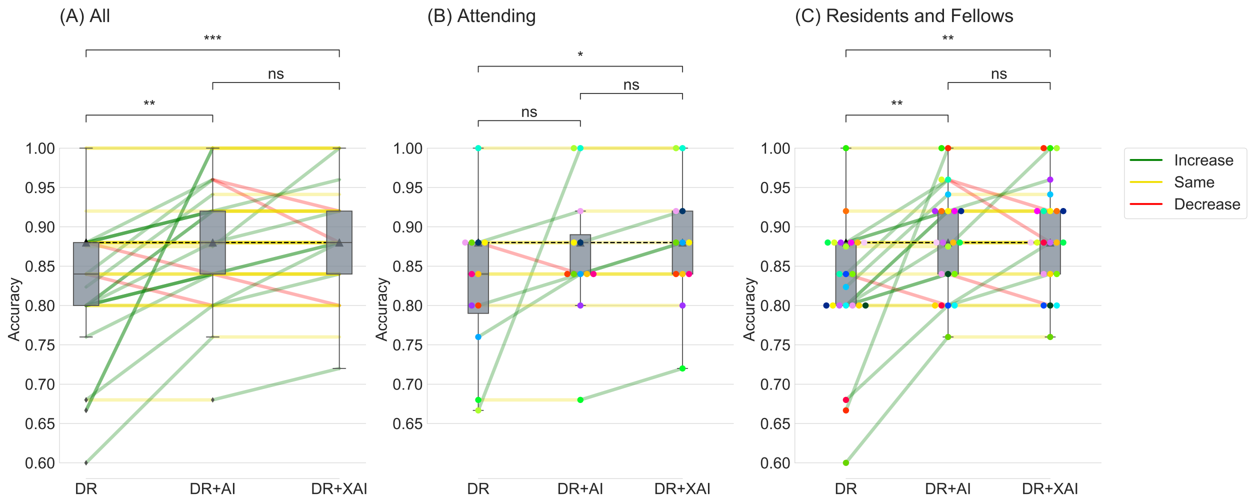

Figure 4: Participants' task performance on glioma grading in three conditions: 1) DR: Physicians performing the task alone; 2) DR+AI: Physicians performing the task with AI assistance (with predictions from AI); 3) DR+XAI: Physician performing the task with XAI assistance (with predictions and explanations from AI). All participants' data are visualized in panel (A), and attending physicians' or resident + fellow physicians' data are shown in panel (B) and (C) respectively. For each panel, we show box plots for the three conditions, with lines and dots indicating the change of performance for each participant. The horizontal dashed line indicates AI accuracy of 0.88. The color of the dots indicates each individual participant's performances. The lines connecting the same color of dots indicate a participant's performance change in between different conditions, with green indicating increase, yellow indicating the same, and red indicating decrease. ns: $p > 0.05$, $* : .01 \leq p \leq .05$, $** : .001 \leq p \leq .01$, $*** : .0001 \leq p \leq .001$.

For the accuracy of each condition in the attending physician subgroup, the non-parametric Friedman test showed a statistically significant difference in task accuracies among the three conditions, $\chi^2_F(2) = 8.27, p = .016$. We then conducted post-hoc analysis using Wilcoxon signed-rank tests with Bonferroni correction. The results showed that there was not a statistically different accuracy between the **DR+AI** and **DR** conditions ($Z = 9.5, p = .51$), and between **DR+XAI** and **DR+AI** conditions ($Z = 0.0, p = .14$). However, the **DR+XAI** condition had a statistically higher accuracy compared to the **DR** condition ($Z = 0.0, p = .047$). We also calculated the effect size using common language effect size, and results showed a physician has a probability of 60.8% of having a higher accuracy when assisted by AI prediction (**DR+AI**) than performing the task alone (**DR**), a probability of 66.7% of having a higher accuracy when assisted by AI prediction and explanation (**DR+XAI**) than performing the task alone (**DR**), but only a probability of 56.3% of having a higher accuracy when assisted by AI prediction and explanation (**DR+XAI**) than assisted by AI prediction alone (**DR+AI**).

| Condition | N | M±SD | Min | 25% Q | Mdn | 75% Q | Max |
|---|---|---|---|---|---|---|---|
| **DR** | 23 | $82.46 \pm 8.58$ | 60.00 | 80.00 | 84.00 | 88.00 | 100.00 |
| **DR+AI** | 23 | $88.42 \pm 6.67$ | 76.00 | 84.00 | 88.00 | 92.00 | 100.00 |
| **DR+XAI** | 23 | $88.96 \pm 6.66$ | 76.00 | 84.00 | 88.00 | 92.00 | 100.00 |

Table 5: Descriptive statistics for resident and fellow physicians' task performance accuracy (%). N - number of participants, M - mean, SD - standard deviation, Q - quantile, Mdn - median.

For the accuracies in each condition in the resident and fellow physician subgroup, the non-parametric Friedman test showed a statistically significant difference in task accuracies among the three conditions, $\chi^2_F(2) = 16.98, p = .0002$. We then conducted post-hoc analysis using Wilcoxon signed-rank tests with Bonferroni correction. The results showed that the **DR+AI** condition had a statistically higher accuracy compared to the **DR** condition ($Z = 2.0, p = .004$); similarly, the **DR+XAI** condition had a statistically higher accuracy compared to the **DR** condition ($Z = 2.0, p = .004$). However, the accuracies between **DR+AI** and **DR+XAI** conditions did not show statistically significant difference ($Z = 10.5, p = 1.6$). We also calculated the effect size using common language effect size, and results showed a physician has a probability of 70.2% of having a higher accuracy when assisted by AI prediction (**DR+AI**) than performing the task alone (**DR**), a probability of 73.2% of having a higher accuracy when assisted by AI prediction and explanation (**DR+XAI**) than performing the task alone (**DR**), but only a probability of 52.4% of having a higher accuracy when assisted by AI prediction and explanation (**DR+XAI**) than assisted by AI prediction alone (**DR+AI**).

To visualize the change of participants' task performance of participants in each condition, we additionally visualize the receiver operating characteristic (ROC) curve (upper row) and precision-recall (PR) curve (lower row) in Fig. 5 for AI and participants in each condition. The performance change as indicated by the arrows showed participants' performances had the tendency to pointing to the direction of better performance area (for ROC plot, it is the upper left corner; for PR plot, the upper right corner). And most of the arrow heads landed on areas on or above AI's curve, indicating physicians' performance with AI assistance is equivalent or better than AI's performance alone. This aligns with the main finding that AI assistance improved physicians' task performance. And such performance boosting is more prominent in participants whose initial task performance is inferior to AI performance (below the AI ROC or PR curve).

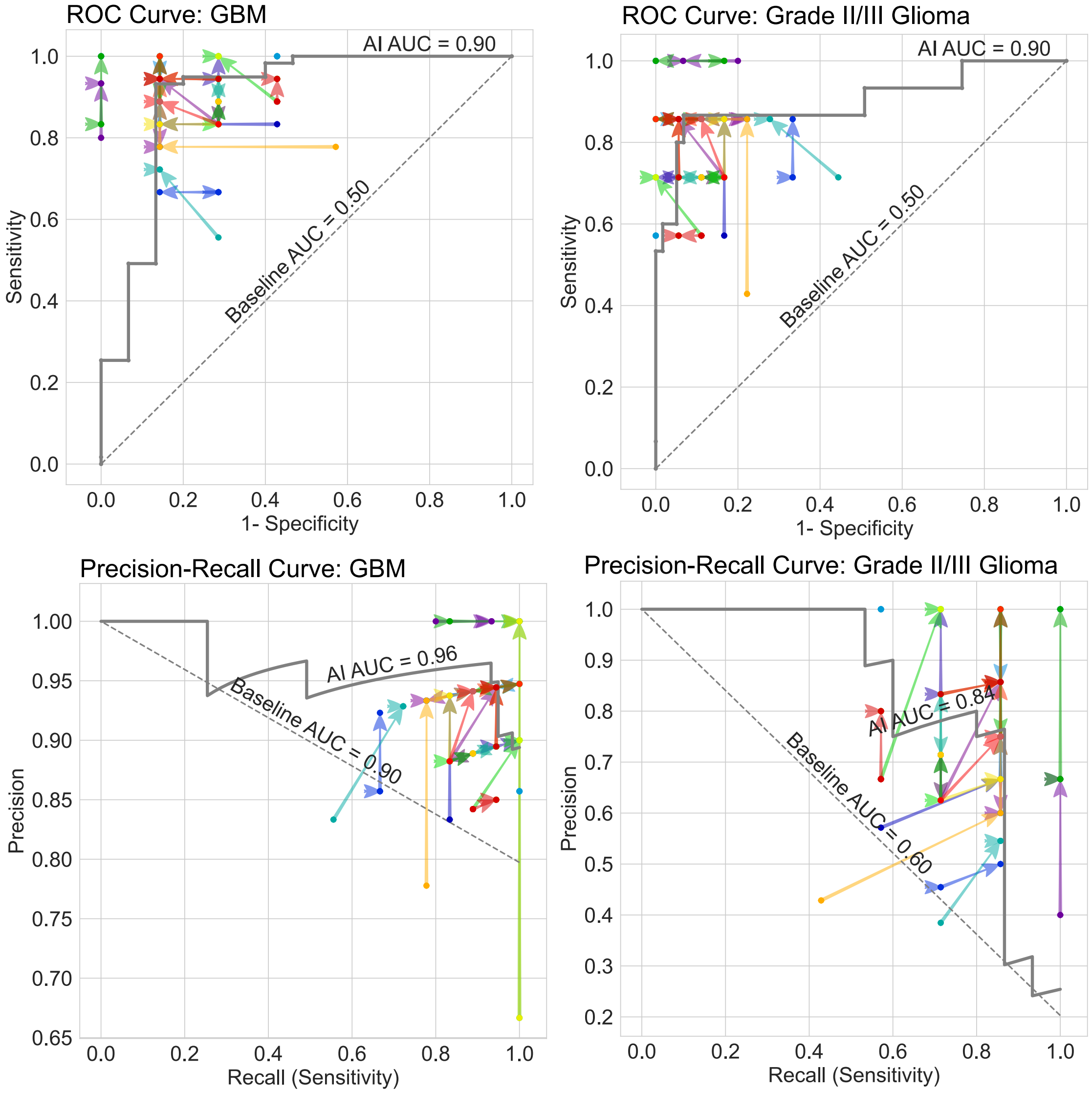

Figure 5: The receiver operating characteristic (ROC) curve (upper row) and precision-recall (PR) curve (lower row) to evaluate the performance on GBM (left) and grade II/III glioma prediction (right). In each plot, the AI model's performance is indicated as the gray curve, and the physicians' performance is indicated as dots, with different colors representing different participants. The arrows in between dots are the performance change between conditions. For ROC curve, a better performance would be near the upper left corner. For PR curve, a better performance would be near the upper right corner. We also indicate the AUC (area under the curve) value for the curve of AI and random guess baselines (the dashed gray line). Each color indicates a particular participant, and the two arrows per color are: from **DR** (doctor performing the task alone) to **DR+AI** (doctor assisted by AI), and from **DR+AI** to **DR+XAI** (doctor assisted by XAI).

We also computed the correlation between physicians' task performance (measured using accuracy) improvement and their clinical experience, using Pearson's r correlation coefficient. The results showed that there was negative moderate correlation ($r = -0.32$) between physicians' years of practicing neurosurgery, and their performance improvement after AI prediction assistance (the performance difference between **DR+AI** and **DR**). This indicates physicians' clinical experience may be an indicative factor to predict AI assistant performance improvement, with junior physicians benefiting more from AI prediction assistance.

Furthermore, there was negligible correlation ($r = -0.15$) between physicians' years of practicing neurosurgery, and their performance improvement after AI prediction and explanation assistance (the performance difference between **DR+XAI** and **DR**). This may indicate physicians' performance improvement with both AI prediction and explanation assistance may be an even more complex process, and physicians' clinical experience may not be a good single factor to predict the outcome of this process. We plot the correlation regression lines in Fig. 6.

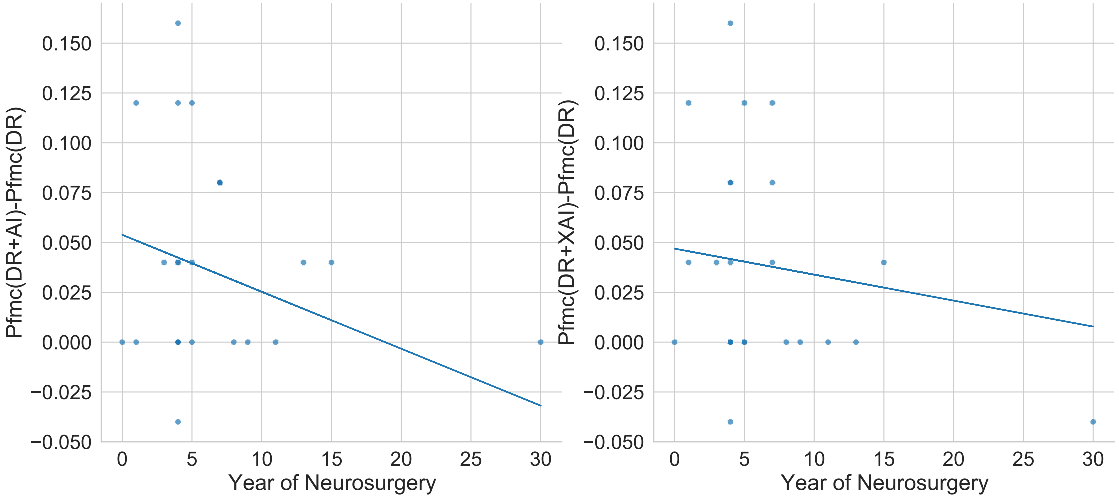

Figure 6: Scatter plot between participants' years of practicing neurosurgery, and their accuracy improvement with the assistance of AI prediction (left), and with the assistance of AI prediction and explanation (right). We also plot the regression lines for each plot, and the coefficients of the regression lines are -0.003 and -0.001, respectively.

#### 2.3 Decision agreement and decision change

For the decision agreement pattern of the subgroups of attending vs. resident+fellow physicians, as shown in Fig. 7, as a baseline when physicians performed the task alone (**DR** condition), the decision agreement is 82.7% (210) for attending physicians, and 80.2% (405) for resident+fellow physicians. When physicians were assisted by AI's prediction (**DR+AI** condition), the decision agreement increased to 85.4% (217) for attending physicians, and 87.5% (442) for resident+fellow physicians. When physicians were assisted by AI's prediction and explanation (**DR+XAI** condition), the decision agreement is the same, 85.4% (217) for attending physicians, and slightly increased to 88.1% (445) for resident+fellow physicians.

The physicians' subgroup decision change patterns have similar trends as analyzed in the manuscript, shown in Fig. 7.

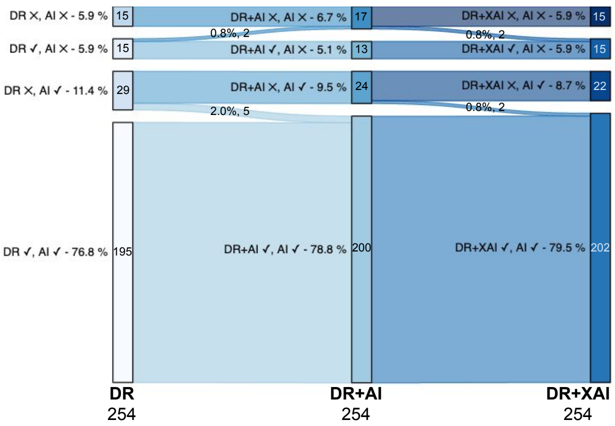

Figure 7: Participants' decision change stream plot for each error category for attending physicians (left), and resident+fellow physicians (right). The three columns represents the three conditions of DR, DR+AI, and DR+XAI, respectively. The rectangles in each column show the doctor and AI's decision correctness, and the decision agreement is the first and fourth rectangles, where doctor and AI made the same decisions (both are correct or incorrect). The percentage and absolute number for each category are indicated. The total number of decisions is 254 and 505 for attending and resident+fellow physicians, respectively.

#### 2.4 Trust and willingness to use AI

For participants' level of trust and willingness to use AI, we conducted fine-grained subgroup analysis based on two subgroups of participants' clinical positions: 1) attending physicians; 2) resident and fellow physicians. We used the non-parametric Friedman test to test if there are significant difference of participants' trust and willingness to use AI at three time points: 1) the initial baseline without knowing any information from AI; 2) after viewing AI's performance metrics, and 3) after using AI's predictions and explanations for the 25 MRIs.

For attending physicians, results did not show a statistically significant difference among the three time points for both trust in AI ($\chi^2_F(2) = 5.55, p = .062$), and willingness to use AI ($\chi^2_F(2) = 1.75, p = .416$). The descriptive statistics are shown in Table 6 and Fig. 8.

| | Time point | N | M±SD | Min | 25% Q | Mdn | 75% Q | Max |
|---|---|---|---|---|---|---|---|---|
| Trust | Bsl | 10 | $5.20 \pm 2.35$ | 2.00 | 3.50 | 5.00 | 5.75 | 9.00 |
| | Pfm | 10 | $6.80 \pm 2.15$ | 3.00 | 7.00 | 7.00 | 8.00 | 9.00 |
| | Use | 10 | $6.50 \pm 2.84$ | 2.00 | 3.75 | 7.50 | 9.00 | 9.00 |
| Willingness | Bsl | 10 | $4.00 \pm 3.40$ | 0.00 | 2.00 | 3.00 | 4.50 | 10.00 |
| | Pfm | 10 | $4.90 \pm 3.28$ | 0.00 | 2.50 | 5.00 | 7.50 | 10.00 |
| | Use | 10 | $3.80 \pm 3.39$ | 0.00 | 1.00 | 2.50 | 7.50 | 8.00 |

Table 6: Descriptive statistics for attending physicians' trust and willingness to use AI. N - number of participants, M - mean, SD - standard deviation, Q - quantile, Mdn - median. The three time points are: 1) Bsl: the initial baseline without knowing any information from AI; 2) Pfm: after viewing AI performance metrics, and 3) Use: after using AI's predictions and explanations for the 25 MRIs.

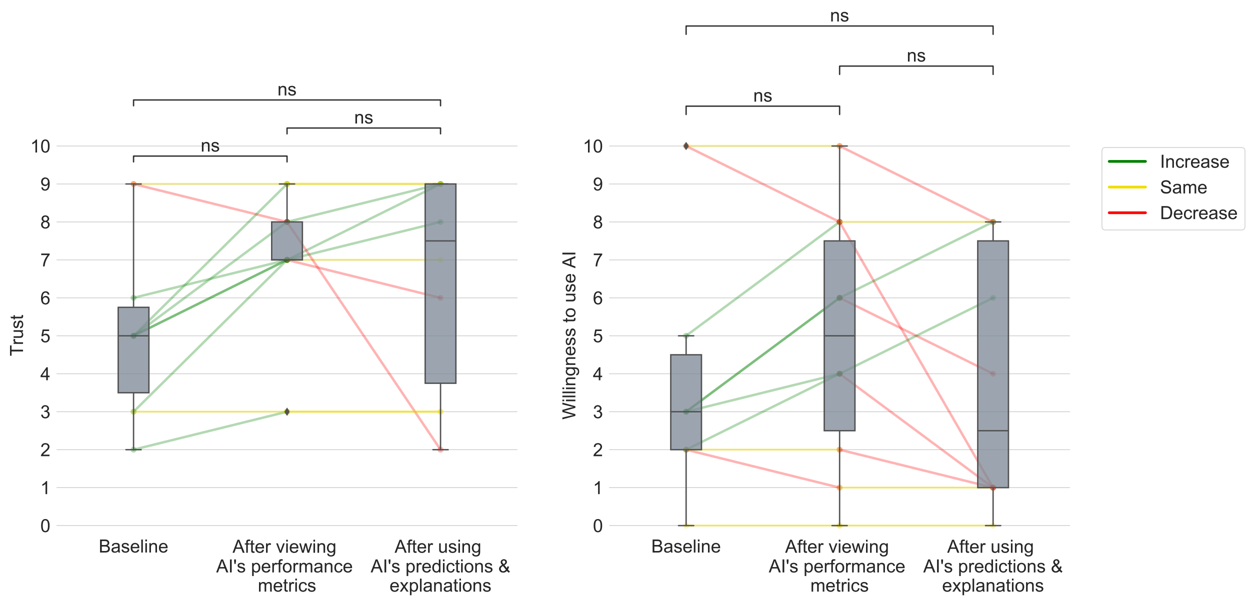

Figure 8: Box plots and changes of attending physicians' trust in the AI (left), and willingness to use AI (right) at the initial baseline, after viewing AI's performance metrics, and after using the AI's predictions and explanations. Both dependent variables are reported on a 0-10 point scale. The lines in between two time points indicate the change for each participant, with green indicating an increment, yellow indicating no change, and red indicating a decrement. ns: $p > 0.05$, $*: .01 \leq p \leq .05$, $**: .001 \leq p \leq .01$, $***: .0001 \leq p \leq .001$.

For resident and fellow physicians, results showed a statistically significant difference among the three time points for both trust in AI ($\chi_F^2(2) = 11.47, p = .003$), and willingness to use AI ($\chi_F^2(2) = 7.86, p = .0196$).

We conducted post-hoc analysis using Wilcoxon signed-rank tests with Bonferroni correction to identify the statistically different pairs. For the level of trust in the AI system, resident and fellow physicians rated a statistically higher trust after viewing AI's performance metrics compared with the initial baseline ($Z = 0.0, p = .00587$); but the trust level did not show statistically significant difference after using AI's predictions and explanations for the 25 MRIs compared with the initial baseline ($Z = 35.5, p = .085$); and there was no statistically difference between the trust level after viewing AI's performance metrics and after using AI's predictions and explanations for the 25 MRIs ($Z = 53.0, p = 2.06$). Similarly for the level of willingness to use AI, participants only rated a statistically higher willingness to use AI after viewing AI's performance metrics compared with the initial baseline ($Z = 9.0, p = .026$); and the rest pairwise test did not show a statistically significant difference. The descriptive statistics are shown in Table 7, and the statistical test results are visualized in Fig. 9.

| | Time point | N | M±SD | Min | 25% Q | Mdn | 75% Q | Max |
|---|---|---|---|---|---|---|---|---|
| Trust | Bsl | 19 | $5.37 \pm 1.92$ | 0.00 | 5.00 | 5.00 | 7.00 | 8.00 |
| | Pfm | 19 | $6.68 \pm 1.42$ | 4.00 | 5.00 | 7.00 | 8.00 | 8.00 |
| | Use | 19 | $6.68 \pm 2.67$ | 0.00 | 7.00 | 8.00 | 8.00 | 9.00 |
| Willingness | Bsl | 19 | $4.16 \pm 2.52$ | 0.00 | 3.00 | 5.00 | 5.00 | 8.00 |
| | Pfm | 19 | $5.16 \pm 1.98$ | 1.00 | 3.50 | 5.00 | 6.50 | 8.00 |
| | Use | 19 | $5.00 \pm 3.04$ | 0.00 | 2.50 | 5.00 | 7.50 | 9.00 |

Table 7: Descriptive statistics for resident and fellow physicians' trust and willingness to use AI. N - number of participants, M - mean, SD - standard deviation, Q - quantile, Mdn - median. The three time points are: 1) Bsl: the initial baseline without knowing any information from AI; 2) Pfm: after viewing AI performance metrics, and 3) Use: after using AI's predictions and explanations for the 25 MRIs.

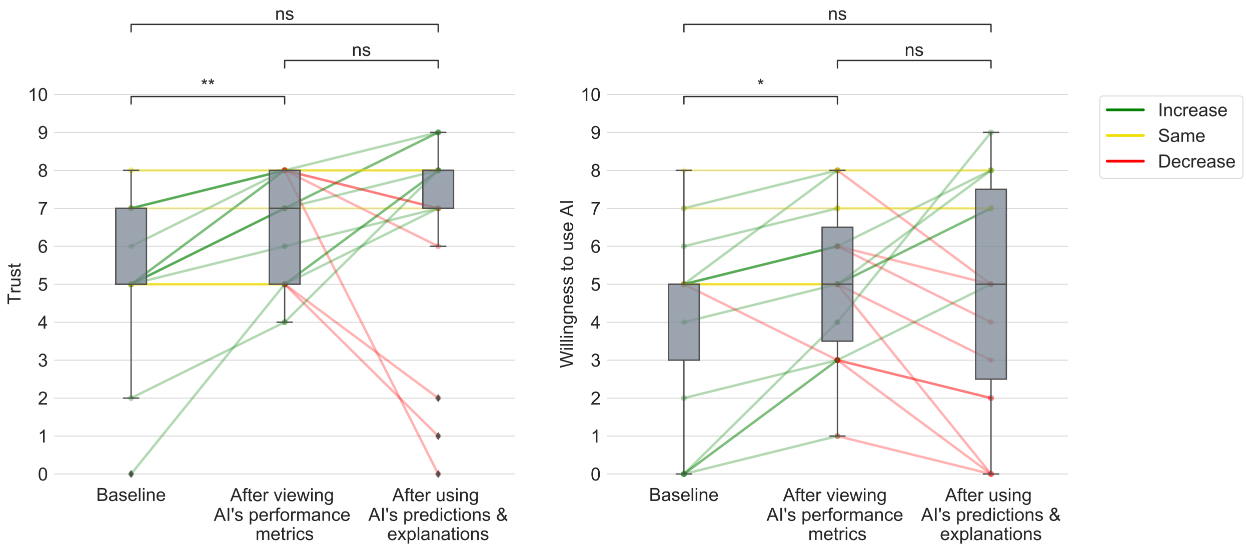

Figure 9: Box plots and changes of resident and fellow physicians' trust in the AI (left), and willingness to use AI (right) at the initial baseline, after viewing AI's performance metrics, and after using the AI's predictions and explanations. Both dependent variables are reported on a 0-10 point scale. The lines in between two time points indicate the change for each participant, with green indicating an increment, yellow indicating no change, and red indicating a decrement. ns: $p > 0.05$, $* : .01 \leq p \leq .05$, $** : .001 \leq p \leq .01$, $* * * : .0001 \leq p \leq .001$.

#### 2.5 Clinical usage scenarios for AI explanation

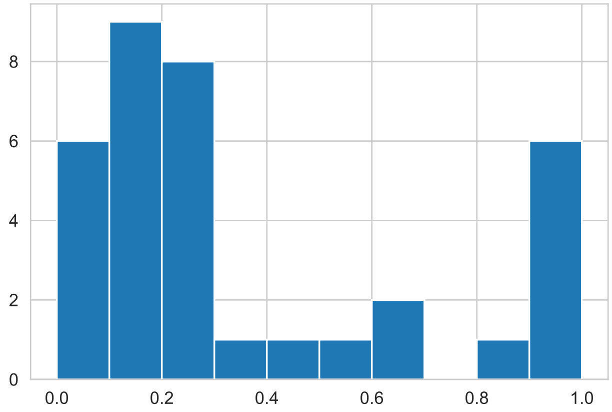

Figure 10: A histogram showing the distribution of 35 participants' need explanation degree. On the $x$-axis, 1.0 indicates the participant need explanation for all the MRI cases, and 0.0 indicates for none of the MRI cases.

[5] Weina Jin, Xiaoxiao Li, Mostafa Fatehi, and Ghassan Hamarneh. Guidelines and evaluation of clinical explainable ai in medical image analysis. *Medical Image Analysis*, page 102684, 2022.

[6] Weina Jin, Xiaoxiao Li, and Ghassan Hamarneh. Evaluating explainable ai on a multi-modal medical imaging task: Can existing algorithms fulfill clinical requirements? *Proceedings of the AAAI Conference on Artificial Intelligence*, 36(11):11945–11953, June 2022.

[7] Scott M Lundberg and Su-In Lee. A unified approach to interpreting model predictions. In I. Guyon, U. V. Luxburg, S. Bengio, H. Wallach, R. Fergus, S. Vishwanathan, and R. Garnett, editors, *Advances in Neural Information Processing Systems*, volume 30. Curran Associates, Inc., 2017.

[8] Marco Tulio Ribeiro, Sameer Singh, and Carlos Guestrin. "Why Should I Trust You?": Explaining the Predictions of Any Classifier. In *Proceedings of the 22nd ACM SIGKDD International Conference on Knowledge Discovery and Data Mining - KDD '16*, pages 1135–1144, New York, New York, USA, 2016. ACM Press.

[9] R. R. Selvaraju, M. Cogswell, A. Das, R. Vedantam, D. Parikh, and D. Batra. Grad-cam: Visual explanations from deep networks via gradient-based localization. In *2017 IEEE International Conference on Computer Vision (ICCV)*, pages 618–626, 2017.

[10] Avanti Shrikumar, Peyton Greenside, and Anshul Kundaje. Learning important features through propagating activation differences. In *Proceedings of the 34th International Conference on Machine Learning - Volume 70*, ICML'17, page 3145–3153. JMLR.org, 2017.

[11] Avanti Shrikumar, Peyton Greenside, Anna Shcherbina, and Anshul Kundaje. Not just a black box: Learning important features through propagating activation differences, 2017.

[12] Karen Simonyan, Andrea Vedaldi, and Andrew Zisserman. Deep inside convolutional networks: Visualising image classification models and saliency maps, 2014.

[13] Karen Simonyan and Andrew Zisserman. Very deep convolutional networks for large-scale image recognition. In Yoshua Bengio and Yann LeCun, editors, *3rd International Conference on Learning Representations, ICLR 2015, San Diego, CA, USA, May 7-9, 2015, Conference Track Proceedings*, 2015.

[14] Daniel Smilkov, Nikhil Thorat, Been Kim, Fernanda Viégas, and Martin Wattenberg. Smoothgrad: removing noise by adding noise, 2017.

[15] Jost Tobias Springenberg, Alexey Dosovitskiy, Thomas Brox, and Martin Riedmiller. Striving for simplicity: The all convolutional net, 2015.

[16] Mukund Sundararajan, Ankur Taly, and Qiqi Yan. Axiomatic attribution for deep networks. In *Proceedings of the 34th International Conference on Machine Learning - Volume 70*, ICML'17, page 3319–3328. JMLR.org, 2017.

[17] Matthew D. Zeiler and Rob Fergus. Visualizing and understanding convolutional networks. In David Fleet, Tomas Pajdla, Bernt Schiele, and Tinne Tuytelaars, editors, *Computer Vision – ECCV 2014*, pages 818–833, Cham, 2014. Springer International Publishing.

[18] Luisa M. Zintgraf, Taco S. Cohen, Tameem Adel, and Max Welling. Visualizing deep neural network decisions: Prediction difference analysis. In *5th International Conference on Learning Representations, ICLR 2017, Toulon, France, April 24-26, 2017, Conference Track Proceedings*. OpenReview.net, 2017.
